# Population-wide analysis of laboratory tests to assess seasonal variation and the relevance of temporal reference interval modification

**DOI:** 10.1101/2022.11.17.22282394

**Authors:** Victorine P. Muse, Alejandro Aguayo-Orozco, Sedrah B. Balaganeshan, Søren Brunak

## Abstract

We identified mortality-, age-, and sex-associated differences in relation to reference intervals (RI) for laboratory tests in population-wide data from nearly two million hospital patients in Denmark and comprising of more than 300 million measurements. A low-parameter mathematical wave-based modification method was developed to adjust for dietary and environment influences during the year. The resulting mathematical fit allowed for improved association rates between re-classified abnormal laboratory tests, patient diagnoses and mortality. The study highlights the need for seasonally modified RIs and presents an approach that has the potential to reduce over- and underdiagnosis, impacting both physician-patient interactions and EHR research as a whole.

## Introduction

Seasonal variation in electronic patient record data (e.g., diagnoses, laboratory values, prescription use) is well documented within many countries and throughout both hemispheres^1,2^. Most investigations, however, have focused on specific disease areas or on their variable mortality rates^2–6^. Investigations of specific laboratory tests are correspondingly focused to these areas, thereby limiting the breadth of research findings for naturally occurring biochemical seasonality^2–6^. A population-wide, comprehensive seasonality study across diseases and their corresponding sets of laboratory tests has yet to be conducted.

Reference intervals (RI) calculated from tests on healthy individuals are universally used in healthcare following International Federation of Clinical Chemistry (IFCC) guidelines^7,8^. RIs are crucial-in patient care by enabling clinicians to interpret laboratory data accurately and combining the information to clinical action. Modern reference intervals do not normally factor in seasonal changes. These limitations, in turn, result in ill-defined reference intervals for defining points of action for patient-physician interactions^9^. It is, therefore, essential to identify these natural fluctuations in laboratory values and, where indicated, adjust intervention protocols accordingly.

Several laboratory values are well documented for their seasonal variation, e.g., vitamin D, folate, and immune biomarkers such as C-Reactive Protein (CRP), neutrophils, and monocytes^2,5,10–13^. The first two can be associated with seasonal changes in daylight hours and seasonal dietary changes, respectively, while the latter list correlates to seasonal allergen fluctuations as well as seasonal disease trends (e.g., pneumonia and flu) ^5,10,11,14–16^. Although this group of tests have been studied repeatedly, they have only been assessed for significance between seasons; no mathematical modelling-based modification methods have been applied to the parameters associated with these fluctuations^16^. The present study accomplishes this task as well as demonstrates its beneficial applications to patient-survival outcomes.

Thyroid Stimulating Hormone (TSH) is also well-documented to have normal seasonal variation shifts, shown to be driven by changes in temperature. A recent study by Wang et al., 2018, emphasizes the importance of redefining reference intervals for such laboratory values since seasonal differences of up to 10% and 15% have been identified^17–19^. These discoveries are crucial in aiding physicians to correctly diagnose thyroid function disorders as opposed to relying on currently used stagnant reference intervals which result in over- and under-diagnoses in thyroid disorders^9,14,15,18,20^.

Another study investigated cardiovascular disease (CVD) and cancer mortality trends in southern vs. northern hemispheres^1^. As expected, CVD followed offset cyclical patterns throughout the year while cancer did not, as cancer – overall – is not known to be a highly seasonal disease^1^. Conversely, coronary vascular disease (CVD) has been known to be seasonally affected by the cold due to its correlation with increased energy expenditure, thereby triggering increased rates of myocardial ischemia and stroke^21,22^. The study suggests that seasonal variation can be specific to a patient’s home hemisphere while non-cyclical values will remain constant throughout the year in both hemispheres despite natural environmental and dietary shifts^1^. Seasonality modelling is also an important tool when studying and predicting new waves of infection during pandemics such as influenza, and now SARS-CoV-2^23^.

While laboratory tests, as noted above, usually have a corresponding reference interval to indicate to the physician whether they are within the normal interval, these intervals do not consider natural and hormonal fluctuations and could therefore misclassify patients^24^.

Further, laboratory tests that do not match a normal fluctuation yet still lie within normal intervals could be more indicative of a patient’s health: for example, a patient with low, yet still normal vitamin D levels during the summer despite spending ample time in the sun^15,18,25^. Of course, the physician in question often has enough experience to determine the abnormality of the given test regardless of the reported reference intervals. On the other hand, Electronic Health Record (EHR) data-driven research cannot – in the same subjective way – account for such assessments and must therefore rely on reported RIs for large scale EHR investigations. These concerns have been raised in other studies^9,26^; however, no comprehensive solution has been proposed.

Several TSH and other hormone-focused studies have made smaller, more limited investigations for the purpose of redefining singular reference intervals^18,19,26,27^. This paper will, instead, make use of all available laboratory tests – presuming enough patient records are available for developing a robust model. As an improvement to these aforementioned studies, we will be able to investigate patient laboratory values over several years and define seasonally adjusted reference intervals for what is considered normal at the local population level, facilitating more accurate physician diagnoses, and assisting in differentiating between disparate subtypes of disease^26,27^. This big data approach has not previously been feasible due to restrictions in access to individual level medical records, making it difficult to accurately define seasonal models across a multitude of tests. Precision medicine approaches may take this type of analysis even further and redefine intervals for each individual^28,29^.

In summary, this study aims to identify seasonality in biochemical tests on a mass scale and propose a method to improve patient test result classification protocols^27,30^. By applying a sinusoidal fit to data such as these, global, big data approaches for identifying seasonally changing parameters can be undertaken. The study further investigates sex, age, and mortality differences in seasonal fluctuations and systematically identify patterns in laboratory tests that can be used to classify patients’ test results more accurately both for patients undergoing clinical work-up and future research investigating the same EHRs.

## Methods

### Data processing and cleaning

Population-wide, in-patient, out-patient, and emergency room data from two Danish Health regions were used in this analysis. A total of 339,609,717 patient laboratory records from the Clinical Laboratory Information System (LABKA) and the Clinical Chemistry Laboratory System (BCC) were collected and systematically processed from the years 2009 to mid-2016, and from 2011 to mid-2016 for each database, respectively. Denmark started a person identification number system in 1968 allowing for continuous and complete record linkage across public hospitals in these regions^31,32^. Private hospital data were not included, as in Denmark these cover less than 1% of the total hospital sector expenditure and often reflect referrals from public hospitals. The impact on the analysis is therefore insignificant [https://www.sundheddanmark.nu/media/1195/sundheddanmark-kort-om-privathospitaler-2018.pdf]. For the purpose of this study, data was further filtered to only include patients whose death status was known at the end of the registry (2016-06-30), thus excluding tourists (0.50%) as to not bias the data towards patients who normally reside in different environments. There is specific coding for tourists as well in the Danish National Patient Registry system (DNPR), also aiding in their removal.

The next stage of data cleaning focused on test codes which largely followed the NPU (Nomenclature, Properties and Units) coding system used in the Nordic countries, including Denmark. The NPU codes are used as identifiers for a given unique clinical laboratory test and the coding structure and development is well defined in a report by Pontet et al.^33^. These codes comprise of the prefix NPU followed by 5 unique digits to identify the test. NPU codes, as opposed to LOINC (Logical Observation Identifiers Names and Codes) codes, better ensure the use of the same code for clinically equivalent tests, making the NPU system the optimal system for this study^34^. Danish hospitals would sometimes use the DNK codes, which have equivalent structure to NPU codes aside from the 3-character prefix, i.e., DNK instead of NPU. They are used when a test does not fit into the NPU terminology [https://kea.au.dk/fileadmin/KEA/filer/Other_reports/119_LABKA_manual.pdf]. Different 3-character prefixes and numeric codes are also sometimes used as hospital specific identifiers^35^. However, employees working with these laboratory systems aided the group with converting codes to the NPU system. A very low portion of tests (0.2%) were not convertible, therefore, for the purpose of this study, languages were conformed to English and unique combinations of source-analyte-unit identifiers that were deemed to be specific enough for direct comparability. After unit corrections, combinations of a unique biochemical analyte and a unique source (i.e., blood, plasma, urine, etc.) were the key criteria for defining a unique test. For example, ALBUMIN – U (albumin in urine) is unique from ALBUMIN – P (albumin from plasma).

Data preprocessing also included several systematic steps to ensure the quality and highest retention of data originally received. A series of lookup dictionaries were created to identify and annotate tests that were incomplete, failed, or otherwise non-readable in the electronic data. Examples of such terminology include “aflyse” (“cancel” in English) and “beregnes ikke” (“not calculated” in English). Iterations of these terms were included as removal terms in the dataset, as the analysis was not actually carried out and was often replaced with a completed one. A mix of Danish and English languages were used in the dataset and therefore the curated dictionaries included both languages. Dictionaries were also used to separate the dataset between qualitative results (positive/negative detection) and quantitative results. For the purposes of this study, only quantitative results were included as we were mainly interested in studying reference intervals and not thresholds for discriminating between positive and negative tests.

Additional cleaning was conducted to ensure uniform formatting of numerical results, for example, aligning all decimal separators to the point system instead of the comma (as is common in Denmark). In addition, character signs such as “+”, “=“ or typos, such as extra spaces and decimal separators, were removed to allow for smooth calculations of quantitative data. Units were converted to one system by correcting for upper- and lower-case discrepancies and deviations in abbreviations used. Lower and upper test reference values from the Danish healthcare system were similarly cleaned up to allow for computational functions to be applied over entire columns of data. These reference intervals were also corrected with supplemental nationally defined reference intervals defined by unique test, age, and sex groups according to IFCC guidelines^8^. Resulting available data included for quantitative processing is summarized in table 1 and visualized in figure 1.

**Table 1:** Data availability overview. This table gives a general statistical overview of cleaned data examined in figure 1a. Results show fairly consistent trends across sexes and regions.

|  | Female | Male | Overall |
| --- | --- | --- | --- |
|  | (N<sub>tot</sub> = ~160M, 51.8%) | (N<sub>tot</sub> = ~150M, 48.2%) | (N<sub>tot</sub> = ~310M, 100%) |
|  | (N<sub>ind</sub> = ~1.06M, 55.2%) | (N<sub>ind</sub> = ~0.86M, 44.8%) | (N<sub>ind</sub> = ~1.92M, 100%) |
| **Age at Time of Test (years)** |  |  |  |
| Mean (SD) | 59.0 (21.8) | 59.9 (20.4) | 59.4 (21.1) |
| Median [Min, Max] | 63.2 [0, 110] | 64.6 [0, 110] | 64.0 [0, 110] |
| **Region** |  |  |  |
| Capitol Region | ~114M (71.0%) | ~106M (71.5%) | 221M (71.2%) |
| Region Zealand | ~46.7M (29.0%) | ~42.5M (28.5%) | 89.3M(28.8%) |
| **Unique Tests per PID** |  |  |  |
| Mean (SD) | 31.5 (20.9) | 31.9 (21.2) | 31.7 (21.1) |
| Median [Min, Max] | 30 [1, 261] | 29 [1, 232] | 29 [1, 261] |
| **Total Tests per PID** |  |  |  |
| Mean (SD) | 151 (311) | 173 (378) | 161 (343) |
| Median [Min, Max] | 56 [1, 16400] | 53 [1, 1900] | 55.0 [1, 1900] |

**Figure 1:**
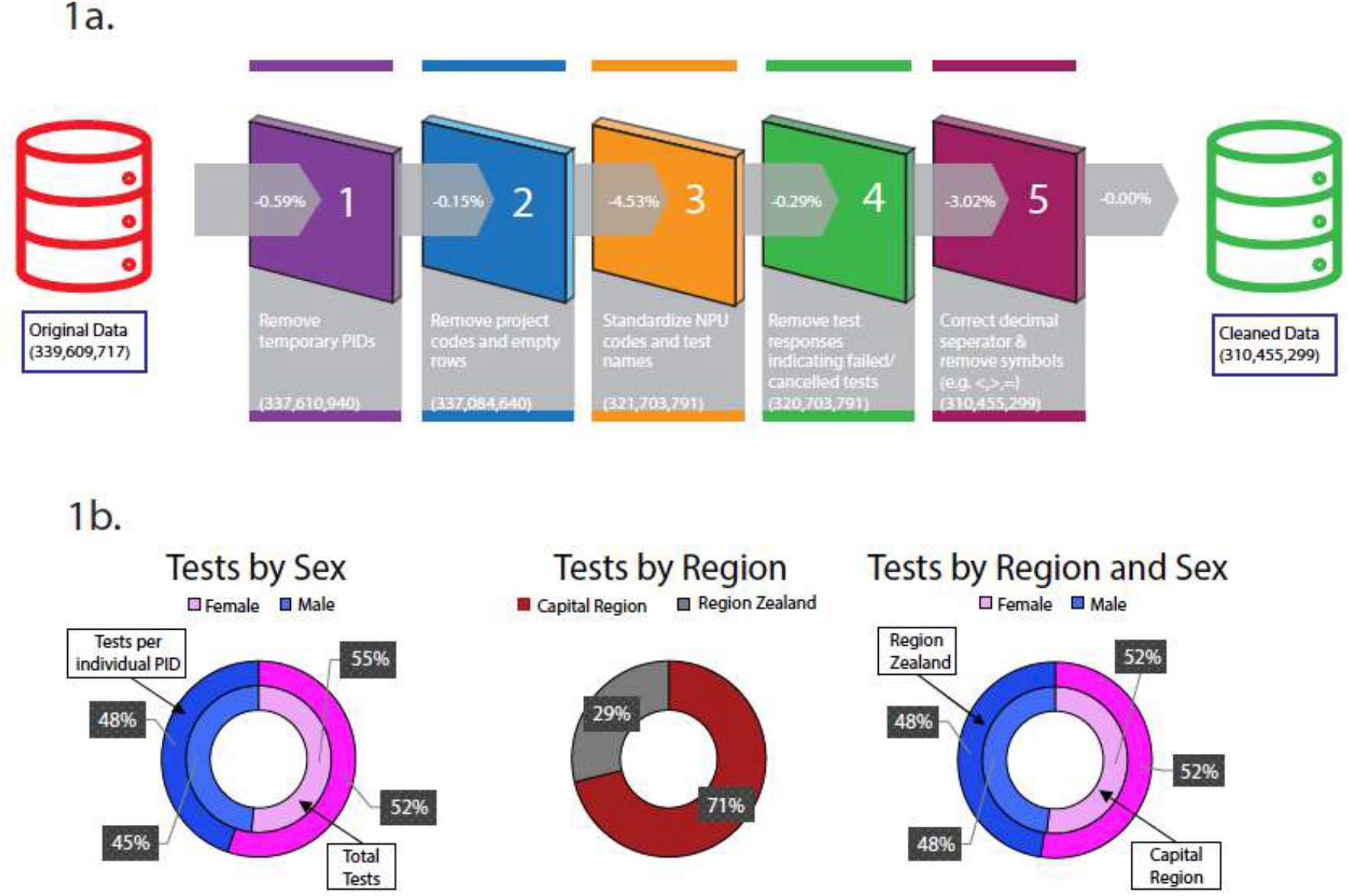
Database cleaning process flow-chart and data overview. Percentage of data removed at each step is indicated as a percentage of the original number of records (339,609,717 observations). Remaining data (310,455,299 observations) is what is then considered cleaned data for use in the research group on a multitude of projects. Figure 1b gives a general overview of sex and location differences within the cleaned dataset. The figures show that men receive more tests per patient on average than women and there is an approximately equal proportion of men and women contributing from each region.

### Normalizing the data to the median

Not all laboratory tests could be assessed for seasonality patterns. The BCC dataset starts later than the LABKA dataset and therefore the combined dataset was a subset of tests from 2012 through 2015 inclusive to allow for yearly complete overviews of data. This step results in 4 complete years of data across both data sets. Mean daily values were used for patients who had repeated measurements of a test in one day; this ensures that a patient can only contribute to the data set once per day per unique laboratory test, as defined above. Differences in units were corrected for, when possible, allowing tests to be more uniform when investigating these unique analyte-source pairs.

Strata for each laboratory test observation were defined by unique combinations of laboratory test, source of biological specimen, unit, lab ID, sex, and age group. Lab ID refers to the location where the test was processed, and it is important to correct tests for this parameter because different labs use different machines or protocols. Additionally, these protocols are frequently updated to meet current scientific standards. Since tests were only studied over a 4-year period, we assumed yearly shifts in normal analyte levels would not be significant. Age groups were defined as 10-year periods. Group 1 included patients aged 0 up to, but not including 10-year-olds; group 2 included 10-year-olds up to, but not including 20-year-olds; and so on. The chosen age group sizes are necessary for age correction in the model; current RI systems often group patients from age 18-65 and therefore the proposed 10-year age groups are already more precise. Patients over 100 years old were excluded from the study as there were not sufficient patients to satisfy stratum requirements. Age group corrections are used to adjust the normalization approach for shifts in normal levels that alter throughout one’s life, as in, for example, during puberty. Given this, seasonality modification for ages below 20 are not as feasible to adjust seasonally due to different starting dates of puberty. As such, age groups 1 and 2 are removed from later parts of the study. Each stratum was required to contain at least 500 unique measurements to ensure a robust and statistically sound dataset. Observations were then normalized to the median value of each of their assigned strata described above. The resulting pre-processed dataset included 279,871,658 unique observations of 421 unique laboratory tests that could then be investigated easily for whole-test seasonality or by sex, age, and mortality differences as described below.

### Sinusoidal model

Using R software (version 4.0.0), data was fit to Equation 1 using a non-linear least-squares algorithm (NLS). This method is commonly used to estimate the parameters of a non-linear model^36^. Parameters were bounded to allow for height and amplitude to float between -1 and 1, and offset to float between 0 and 52. The “port” algorithm was used for fitting as it is the only one from this package that allows for upper and lower bounds to be defined. NA values were omitted from the fit. Weights correlating to the number of available measurements were also included to reduce bias from abnormal hospital visits during the holidays and summer months when only extremely sick people typically go to the hospital.

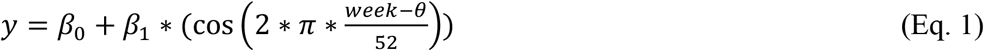

where “y” is the dependent variable that indicates the predicted normalized value for the given test; week is the independent variable for the known week of the calendar year on which the test occurs; “*β*_0_” indicates the height of the function; “*β*_1_” refers to the amplitude of the test; and “*θ*” indicates the offset of the week to shift values seasonally. Figure 2 provides an overview of how the parameters affect the shape of seasonality in each test.

**Figure 2:**
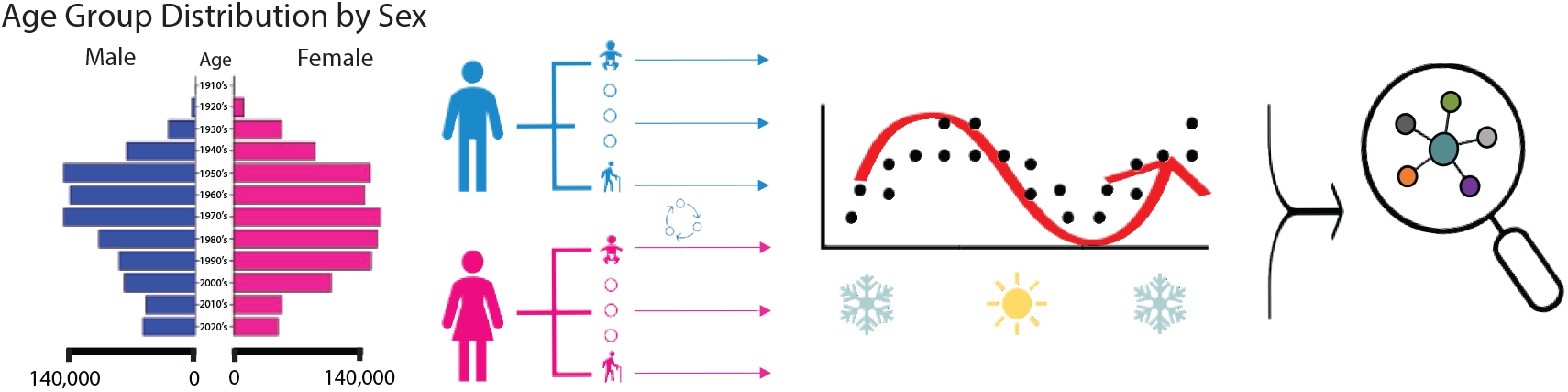
Method flow schematic. This figure demonstrates the main mathematical approach in detecting seasonality in each test. Tests are grouped by sex and age and then fit mathematically to a sinusoidal curve. parameter fits are then investigated using several clustering techniques.

### Checking for seasonality patterns by strata

Tests were compiled by median normalized value per week-year, correcting for weekday vs. weekend variations in testing, year over year shifts changes, as well as daily variation. This approach allows for a single patient to contribute a maximum of 7 times per data point. In order to improve the robustness of the model, a minimum of 100 unique patients were required to include the relevant week-year data point. This additional requirement reduced the viable laboratory tests to 404 resulting in a series of 4, 52 × 404 matrices comprising of weekly median values per unique laboratory test, unit corrected. There will therefore be a maximum of 4 unique data points per week of the year for the mathematical model to be fit to.

This process was repeated for each age, sex, and mortality group and fed through the above-described mathematical model. Outputs were in the form of 3 parameter fits per test and strata and corresponding corrected p-values as reported in supplemental table 1, and visualized in figures 3, 4, and 5. Sex-specific tests are missing from the opposite sex’s final matrix, for example, Prostate Specific Antigen (PSA). Mortality rates were stratified as tests that were taken within 28 days of date-of-death or else patients who survived for more than 28 days, as is standard for mortality studies in clinical medicine.

**Figure 3:**
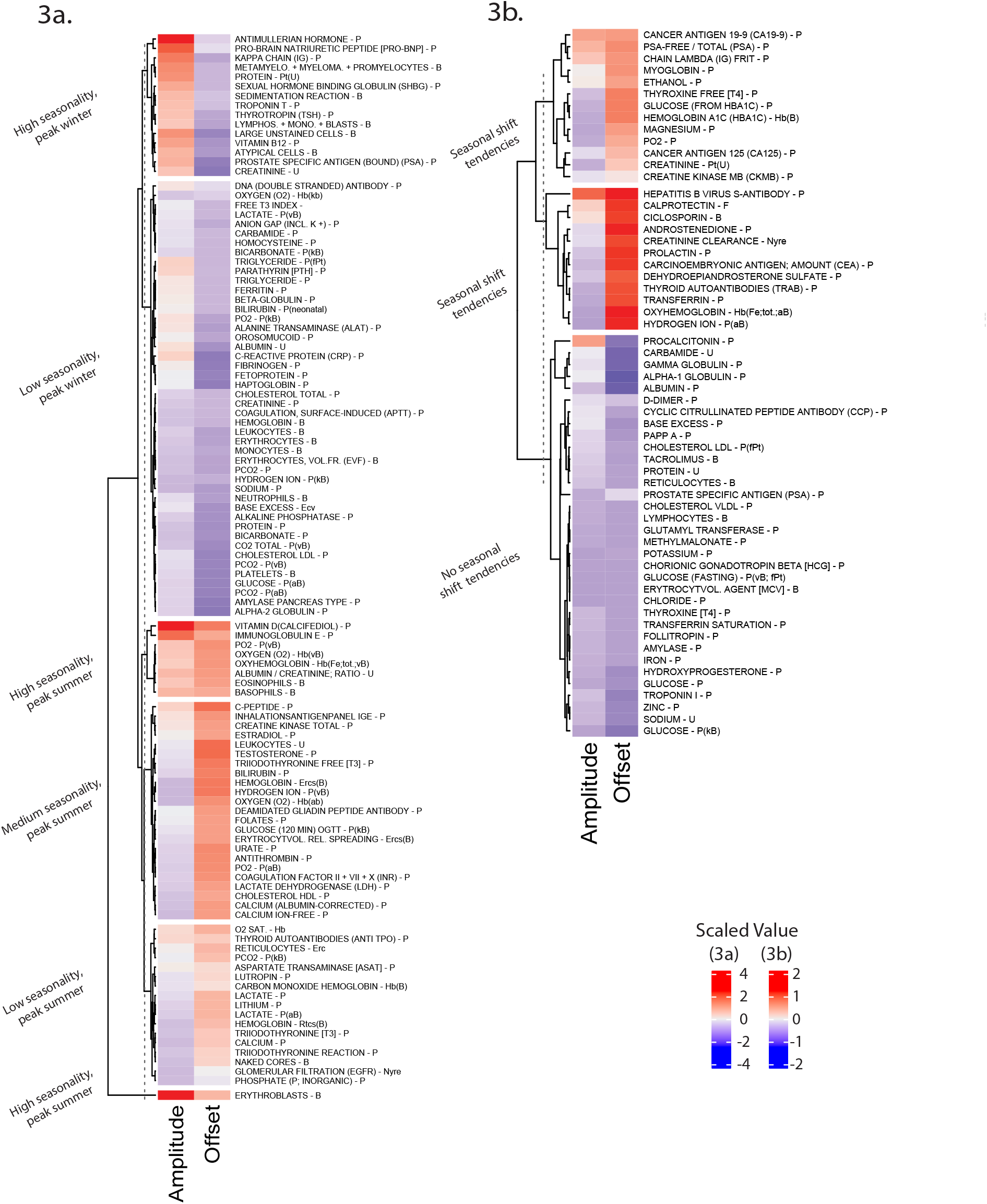
Heatmap for all eligible unique laboratory tests (denoted as analyte-source pair). Figure 3a uses a K-means analysis to cluster tests (with at least one significant amplitude or offset parament fit, FDR corrected pval <0.05) K-means analysis to cluster tests (with at least one significant amplitude or offset parament fit, FDR corrected within 6 groups which have been empirically labelled as seen based on amplitude/offset patterns within the cluster. Figure 3b similarly clusters the remaining, non-significant tests into 3 groups, also empirically labelled based on tendencies seen. Bluer offset values indicate peak values are in the winter while redder values indicate peaks occur in the hottest months. For amplitude fits, higher values correlate to higher amplitude fits while lower values correlated to lower amplitude fits. Parameter values are scaled, but numerical amplitude fits while lower values correlated to lower amplitude fits. Parameter values are scaled, but numerical values are reported in supplemental table 1.

**Figure 4:**
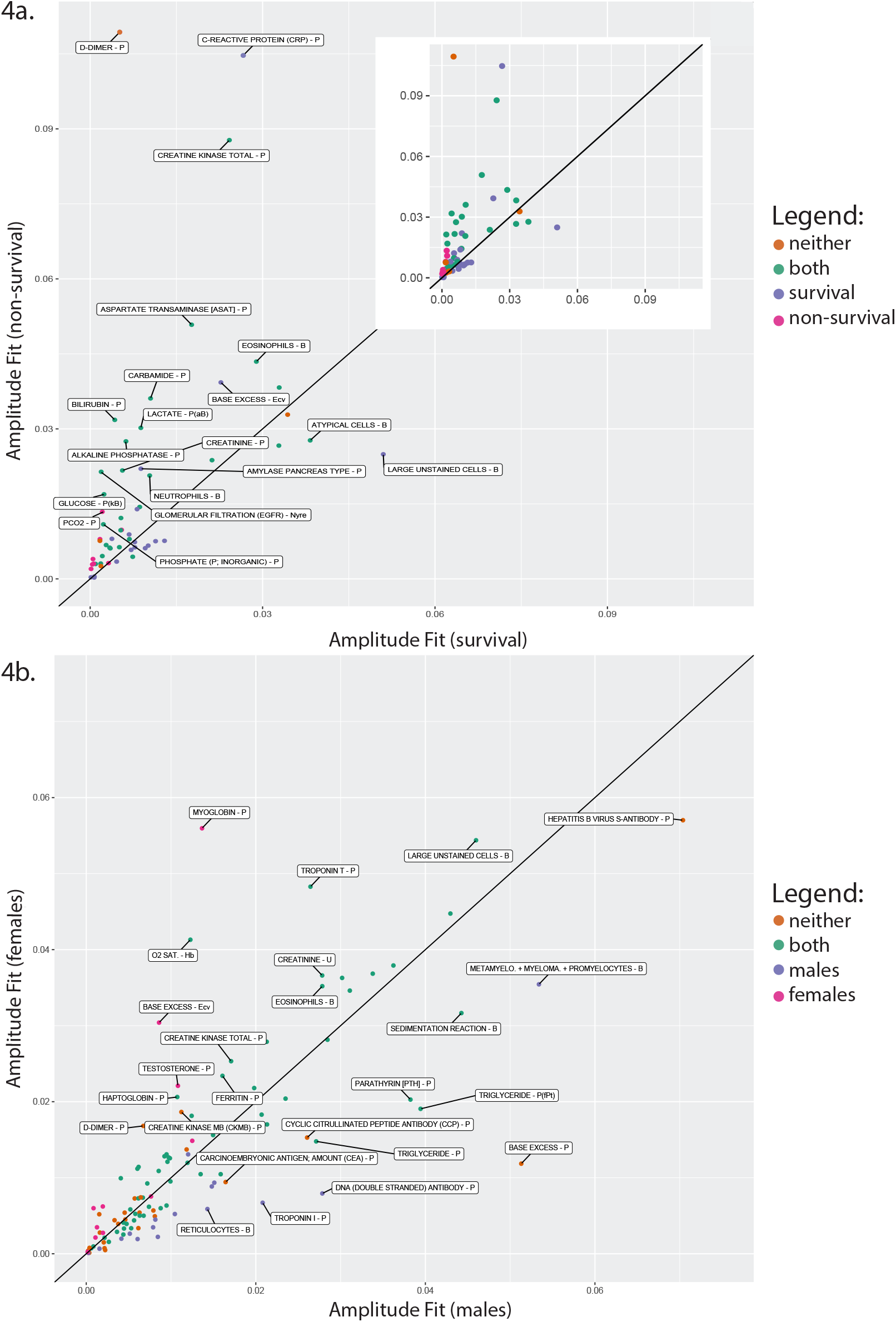
Overview of parameter fits by mortality (figure 4a) and sex (figure 4b). Linear black lines are used in both figures to note where parameters for each stratum would be to be equal. Labels are included for the tests if the differences between the amplitude parameter fits were more than 0.007. Colors indicate whether the parameter was significantly fit for either, both, or neither stratum accordingly (FDR corrected, pval <0.05). Figure 4a includes a second print without the labels to enable an easier view of the color trends. There are 2 data points that were out of range in figure 4b and are therefore listed here: ERYTHROBLASTS – B (0.28, 0.32) and VITAMIN D (CALCIFEDIOL) – P (0.11, 0.08). Numerical values of each data point are provided in supplemental table 1.

**Figure 5:**
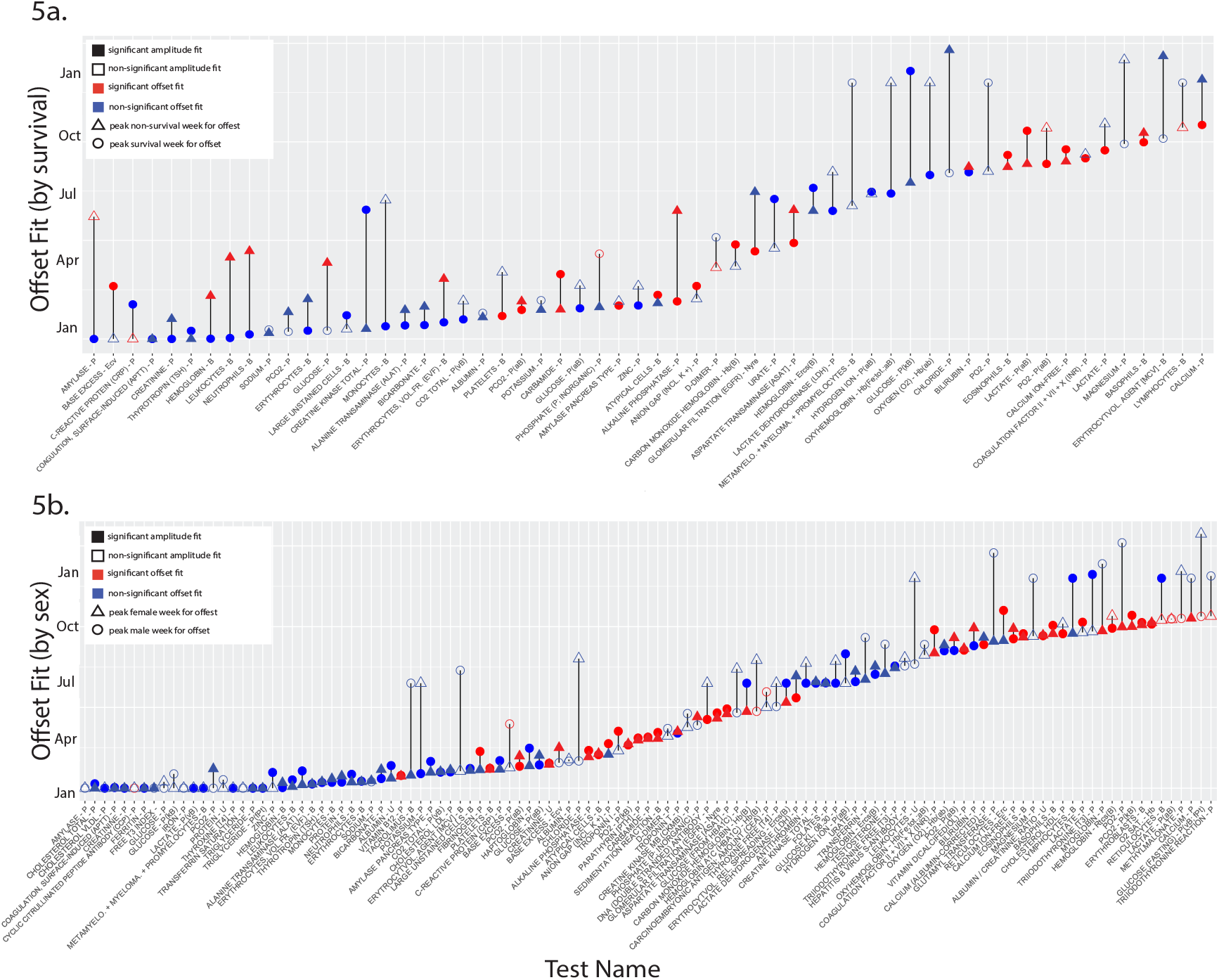
Overview of the offset parameter fits by mortality status (figure 5a) and by sex (figure 5b). Both figures denote where the value peaked by week of the year and whether the fit was significant as denoted by the legend (FDR corrected, pval <0.05). Further information was provided as to whether the correlating amplitude value was significantly fit (FDR corrected, pval<0.05). For both figures, only tests that had available fitted parameters for each stratum were included; those without paired fits, such as for sex-specific tests, are provided in supplemental table 1 as well as all the numerical values used to make this figure.

### Investigating mortality fits

Parameters were fit to the entire preprocessed data set stratified by survival or non-survival using 28 days as the cutoff, sex and age corrected. Amplitude and offset parameter fits were compared more directly in figures 4a and 5a. Significant fits are labelled as such using a pval threshold of <0.05, FDR corrected.

### Investigating sex and age fits

Parameters were first calculated for each age/sex strata individually and examined together as seen in figures 4b and 5b. Larger amplitude values highlighted in figure 4b indicate more seasonality changes in the data. Sex differences were further examined in figure 5b for both amplitude and offset differences, where again significant parameter fits were labelled as such using a threshold of pval <0.05, FDR corrected. Tests with significant (pval < 0.05 FDR corrected) amplitude fits were plotted by age and stratified by gender to explore age and sex differences for each laboratory test in supplemental figure 1. Only tests where both sexes and at least 4 age groups were represented are included. 3D mesh figures of these trends are further visualized in supplemental figure 2 where a minimum of 3 significantly fit age groups were required to be included.

### Adjusting reference intervals with new parameter values

Once a defined curve has been identified, the given equation can be applied to the corresponding known IFCC standard reference value, allowing for fluctuations throughout the year to be visualized. For age groups >2 (reducing bias from puberty) and for all sexes that had significantly fit parameters (listed in figure 6), the corresponding IFCC standard RIs were adjusted using equation 1 and all applicable tests were relabeled as normal or abnormal according to the seasonally adjusted RIs. Operating under the assumption that the new system is the “truth,” tests that changed from normal to abnormal were labelled as false negatives (fn) and those that changed from abnormal to normal were labeled as false positives (fp). All other tests were considered true negatives/positives (tnp) as fitting. This new data set was then integrated with the diagnostic registry, and frequency counts were generated for co-occurrence pairs, where “co-occurrence” is defined as a combination of a fn/fp/tnp laboratory test and a diagnostic code occurring from 2 weeks before the disease code hospital encounter date to 2 days after, as a simple way to look at correlations of testing used for diagnostic purposes. Time to diagnosis can vary greatly by disease type and therefore this timeframe is used for a general overview of trends, not as a validated causal relationship.

**Figure 6:**
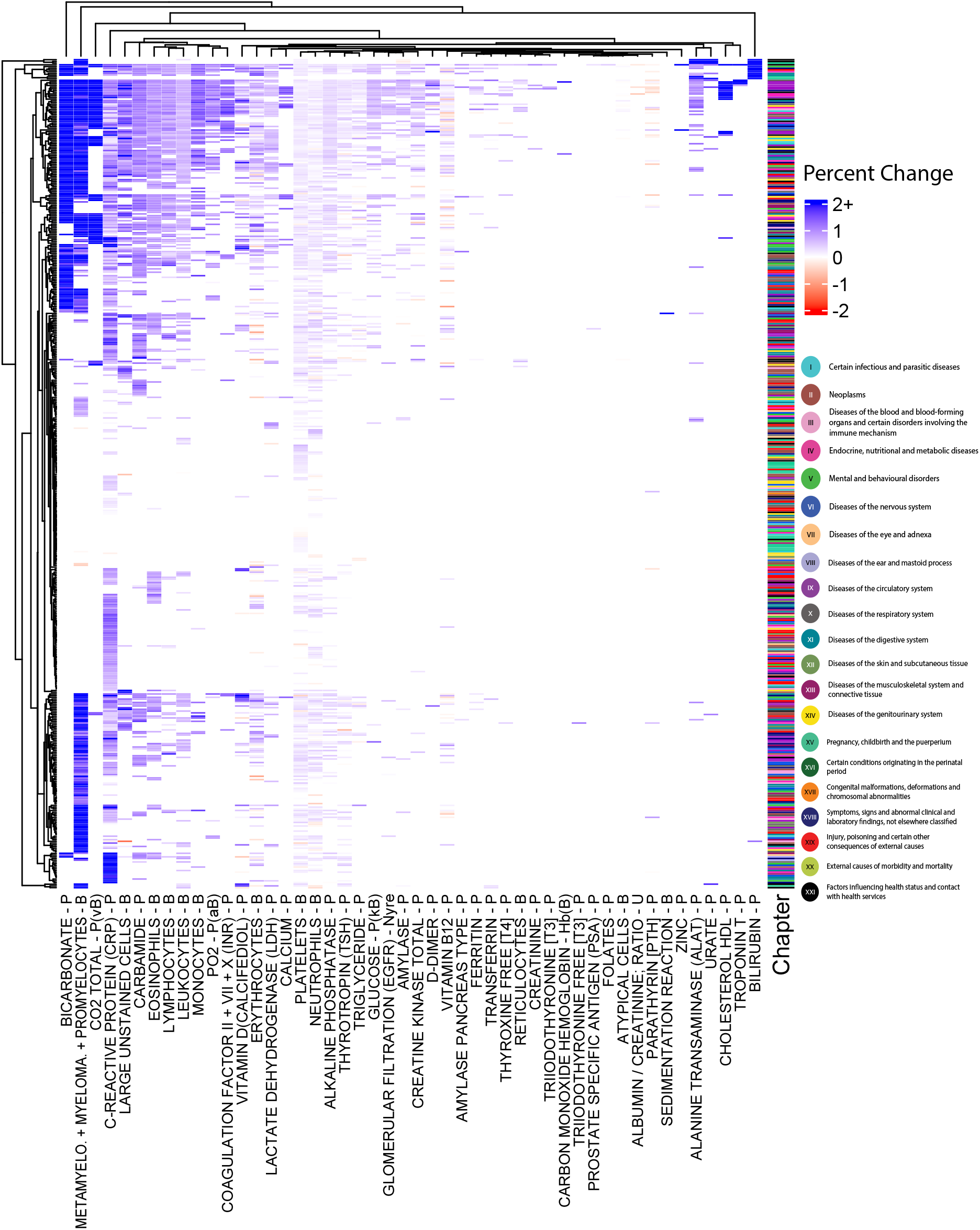
Heatmap of seasonally adjusted model result summary. Seasonally corrected laboratory + level 3 ICD-10 code co-occurrence comparing the seasonally adjusted RI values to the original standard ones. A co-occurrence indicates a laboratory test was labelled as abnormal according to the respective RI system used and the given ICD-10 code was assigned to the same patient. These occurrences only counted if the test in questionoccurred 14 days before the ICD-10 code and up to 2 days after the hospital encounter date. Positive blue bandsindicate a corresponding percentage increase in co-occurrence while red bands indicate a percentage decrease in co-occurrence. Increases in change were often greater than 2%, but to allow for better visualization, anything over 2% is denotated as “2+”; conversely, decreases were never less that -1.5%. White or empty bands indicate that there was insufficient data (based on selection criteria), or there was no prominent change in co-occurrence. Laboratory tests and level 3 ICD-10 codes were allowed to cluster using the hclust algorithm in R.

In order to explore disease patterns, diagnostic codes were shortened to level 3 ICD-10 codes according to the ICD-10 terminology and only pairs that included at least 500 unique patients and 1000 co-occurrences were retained to maintain a more robust overview. Results are seen in figure 6 where, for each unique laboratory test + level 3 ICD-10 code pair, a net percentage increase in fn is shown via heatmap. These were calculated by subtracting the total fp from the total fn occurrences per pair and dividing by the total frequency of co-occurrence and multiplying by 100 to transform them to percentages. Positive percentage values indicate the adjustment increased the pairs association and negative percentage values decreased the association, with respect to the original RI system.

## Results

The original dataset included 339,609,717 laboratory measurements derived from 1,987,180 unique patients in a population-wide setting. It was cleaned as outlined in figure 1a (see also Methods) resulting in 310,455,299 usable unique quantitative measurements for this study. These data are summarized in figure 1b and table 1 where we can see that females, on average, receive less laboratory tests per hospital encounter than males; however, there are more women than men represented in the database. Additionally, there is an equal contribution of men and women to the dataset from Region Zealand and the Capital Region. Approximately 0.5% (1.6 million observations) of the cleaned dataset contained qualitative information such as colors of specimen or genotyping and were excluded from the mathematical analysis.

After filtering for stratum requirements as described in Methods, 1,924,869 unique patients corresponding to 1,545 unique laboratory tests (defined as a unique analyte-specimen pair) were examined for indications of seasonality using the mathematical approach outlined in figure 2. With filtering and requirements for robustness visually described in figure 2, a total of 404 unique laboratory tests were eligible to be assessed for seasonality. Parameter fits and coordinated FDR corrected p-values are reported in supplemental table 1. Minimum data requirements for the algorithm resulted in 169 seasonally fitted laboratory tests, 110 of which have significant amplitude and/or offset shifts (pval <0.05 FDR corrected). Figure 3 summarizes the main seasonality trends for these 169 tests in a heatmap, stratified by significance level. K-means clustering allowed us to further cluster tests by degree of seasonality and by what time of year the value is highest. A Mann-Whitney-Wilcoxon test was performed to confirm there was no significant correlation between the time of day of test taken with whether the given laboratory test was deemed significant (pval = 0.26).

### Mortality stratification

These tests were stratified by mortality risk, i.e., whether the patient in question died within 28 days of testing. Parameter fits comparing mortality risk are reported in figures 4 and 5. Results show significant differences for several tests indicating that for some tests there is a normal seasonality trend in surviving patients while different trends were found in patients who died within 28 days. Standout tests in figure 4a include D-Dimer – P, C-Reactive Protein – P, and Creatine Kinase Total – P. These tests all have higher amplitude fits for non-survival patients than surviving patients which makes sense as they are indicative of very serious disease: blood clotting, pulmonary embolism, deep vein thrombosis, infection/acute inflammation, ischemic heart attack, rhabdomyolysis and more^37–39^. Other tests that have higher amplitude fits for non-surviving patients generally relate to seasonal diseases such as respiratory diseases and infections which are known to be seasonally high during winter. Large unstained cells – B is the main test where surviving patients are significantly higher than their non-surviving counterpart. This test is a marker for viral infection or fungal infection, largely treatable disease categories that should not result in a 28-day mortality risk^40^.

Figure 5a shows the seasonal shift in peaks for each test, stratified by survival or non-survival. This figure clarifies that while both patient groups can experience similar amplitude parameter fits, their peaks occur at different times of the year. A key example of this is for immune response biomarkers where surviving patients will have elevated values during allergy seasons while their non-surviving counterparts experience peaks largely in the colder months due to surges in respiratory diseases.

### Sex stratification

This same stratified approach was completed for males vs. females as seen in figure 4b and 5b. In figure 4b there are several tests where women experience higher amplitude fits than their male counterparts. These tests such as Myoglobin – P, O2 sat. – Hb, haptoglobin – P, and Ferritin – P are all related to iron and hemoglobin binding and oxygenation in the blood. Women are more susceptible to anemia, which is seasonal in itself^41,42^. Additionally, there are diverging trends in males/females in the Troponin I and Troponin T. According to a study by Welsh et al., elevated Troponin T is correlated to more non-CVD related deaths while elevated Troponin I is more closely tied to some CVD outcomes^43^. It is well documented that CVD is seasonal and occurs at higher rates in men, indicating that this finding is also well founded^44^.

Correspondingly, figure 5b shows the different seasonal peaks for men vs. women. They are not as pronounced as in the mortality data, which is reasonable because dietary and environmental shifts should generally match across sexes. A tendency of differences in men and women with thyroid related tests is seen, which is driven more hormonally and therefore sensible, although these offset differences are not statistically significant.

### Sex and age stratification

Those tests that were found to have significant seasonality shifts in supplemental table 1 were then examined for age- and sex-specific shifts in seasonality. These trends are reported in supplemental figure 1 by unique lab test. Amplitude fits reported by sex and age are plotted by age group. The significance of the amplitude fits is noted. While some tests have significant parameter fits overall, they do not translate to all amplitude and offset fits by sex and age group. However, clear trends in age differences can still be observed, such as in vitamin D and TSH. 3D meshes of tests with significant changes shown in supplemental figure 2 revealed some interesting patterns as well. Several tests such as eosinophils, TSH, vitamin D, and vitamin B12 have saddle-plot style trends which emphasizes why more specific seasonality correction by age and sex are so vital to correctly capturing a patient’s health status at the time of testing.

### Improvement by adjusting for seasonality

The overall success of the modification method is shown in figure 6 where patterns of laboratory tests and ICD-10 code pairs can be visualized by increased co-occurrence, decreasing co-occurrence, or unchanged co-occurrence. Due to stratum requirements, only 47 tests of the original 110 significant tests (figure 3a) remain. These tests have the most relation, using frequency counts to ICD-10 codes, and are therefore also the most commonly physician-requested tests, making them the most important to study for seasonality trends.

In figure 6, most of the bands are blue, indicating an increased association between the diagnosis and laboratory test. The red bands indicate the reverse case where there is a decreased association in comparison to the original RIs. For the blue bands, while the method seems to indicate an improved labelling system for differentiation between normal and abnormal tests, this is not always desirable. In some disease groups these changes could be irrelevant, such as with pregnancy-related diseases where patterns in testing relate more to the gestational age of the fetus as opposed to the season of the year. Further, upon close inspection of the results, cancer diagnoses also do not benefit strongly from these seasonal adjustments, as cancer is not inherently seasonal. An interesting pattern emerges as well for chapter 9 ICD-10 codes (circulatory system) where HDL cholesterol and troponin T exhibit a cluster of increased co-occurrence.

Looking at trends by testing we can see a cluster of white blood cell counts (eosinophils, leukocytes, lymphocytes, and monocytes) close to CRP, most likely driven by seasonal respiratory disease patterns and allergens as seen in other studies^5^. This cluster is also adjacent to vitamin D tests, which has been increasingly studied for its role in many diseases such as cancer, metabolic disorders, and many more chronic diseases^45^. We also see platelets and neutrophils – another pair of immune response markers – cluster together. Further work would be required to validate and assess the reasons behind both the disease and test clustering trends.

## Discussion

In this study we identified laboratory tests with previously unknown, or otherwise not well-studied, seasonality patterns, as well as confirmed, well-established seasonality-affected laboratory tests using a low parameter mathematical approach. This study was the first to accomplish this using a big data approach as opposed to focusing on disease specific tests. In addition, we show that this mathematical approach can not only be used to identify seasonality patterns across different strata groups, but also to calculate seasonally adjusted reference intervals using the original IFCC based RI values^8^. Enhancements to associations between laboratory tests and ICD-10 codes exemplified in this study have numerous future applications in mortality predictions and other areas^27^.

Improvements in understanding even small changes in the data is integral to patient triaging in emergency settings where high-risk stroke, cardiac, and acute respiratory diseases can be better identified. CVDs, specifically, can be stratified computationally during the colder months due to the cold’s effect on energy expenditure. Conversely, chronic disease patients with natural biochemical fluctuations can be less worrisome to their physicians as they are likely reactions to allergens in the spring and fall, or energy expenditure changes due to decreased temperatures during the winter.

Figure 6 highlights the need for more standard RIs for cancer and pregnancy, as their afflictions are not as directly influenced by seasonal dietary and weather patterns. These patient cohorts follow other patterns, such as with gestational age for pregnancies. These cases are therefore not likely to be well-captured by the low-parameter mathematical model presented in this study. Instead, more specialized curves would be useful although specific gestational age based RIs are already commonly used. While this global finding on the potential impact seasonally adjusted RIs have on EHR research is compelling, there are several specific noteworthy findings that have more immediate applications to the field.

Tests that experience red bands do not form any clear trends; however, at maximum, only 1.5% of a given test experiences lower co-occurrence while increased co-occurrence rates reach a maximum of 17%, solidifying the total cost-benefit of the adjustment algorithm. Further, a decreased co-occurrence rate may prove to be a beneficial outcome as the test involved does not actually relate to the disease at hand. Figure 6 has several red bands for vitamin B12. While there is no disease pattern associated with these changes, generally the vitamin B12 level is well known to drop seasonally due to dietary changes, so having a low level in the summer is not of concern for most patients, as it will fluctuate back into normal interval^14^.

For several tests, there are significant seasonality trends in the alive/healthy population that disappear from the sick/dying population (figure 4a). This can likely be attributed to the fact that the seasonal patterns for healthy patients are those driven by allergen seasonality or normally fluctuating trends, such as in vitamin B12, vitamin D, and TSH. Conversely, those tests for sick patients near death (i.e., confirmed death within 28 days) are not as influenced by seasonality since most of their tests are abnormal all seasons of the year; for example, in palliative care. The converse trends of significant patterns in dying patients and no seasonality in healthy patients can instead be explained by disease seasonality, such as with respiratory diseases that have peaks in the winter. These tests include D-Dimer, CRP, and Creatine Kinase, all of which have higher amplitude fits for non-surviving patients compared to those surviving for 28+ days. These trends indicate that the tests of most interest for the purpose of identifying normal seasonality fluctuations can be found in the tests where they are only significant for healthy patients or significant for both healthy and sick patients. The other groups are likely driven by disease seasonality or have no clear seasonality trends for sick or healthy patients. This model is therefore capable of stratifying patients who are slightly ill with low mortality risk, from those facing high-risk mortality diseases.

This paper, further, largely investigates sex and age differences across tests and can reflect on how the findings compare to other studies in this field. TSH has largely been discussed to have age-based seasonality changes but there is dispute amongst researchers surrounding sex differences^12,46^. The findings in this paper do not suggest any clear sex-driven seasonality patterns. Vitamin D is also an interesting test to examine because while it is known to vary widely throughout the year, even for healthy people, as seen in supplemental figure 1, it has a declining amplitude (i.e., less variance throughout the year) as a patient ages^10^. It is known that as one ages, the skin is less able to transform vitamin D to a usable form for the body, which would explain why older populations do not experience high peaks of vitamin D in the summer, but rather are consistently low throughout the year with little variance^47^. Examples such as this highlight the need for seasonally adjusted RIs across laboratory tests to identify changes in laboratory values which might indicate a move towards disease at an earlier stage in some individuals compared to the healthy population, based on age and sex specific changes throughout the year. As seen in figures 4b and 5b, this model is already capable of identifying and adjusting for female hormonal-based trends from their age-matched male counterpart’s original RI.

While other studies have shown the promise of deep learning or machine learning for defining personalized reference intervals, our objective is to instead define seasonally adjusted reference intervals applicable to the population level^29^. This approach has shown, as seen in figure 6, that simple population adjustments can help identify key diseases that are falsely correlated to incorrect categorization of normal and abnormal tests, potentially reducing noise, for example, in machine learning (ML) models. This approach can be handled at the individual patient level via physicians who are aware of the individual’s seasonal fluctuations, but EHR approaches may not be as well-informed. With the proposed low parameter population-based seasonal adjustment method, future ML models developed for precision medicine tasks can be greatly improved for disease prediction and prognosis models. Future studies in this area will apply this modification method for just such a task. To note, future model versions will also likely be more mathematically complex as parameter fitting will be based on the physiology and mechanics of the specific disease or tests being studied.

There are several areas where this current model could be improved. Other models, such as the Cohen et al. model, were able to capture a healthy population by removing defined chronic conditions and medications that would bias their results for determining patient-specific lab-test guidelines^29^. Future iterations could make use of such a system, but this study was focused on 28-day mortality as the criterion for normal RI and therefore all tests could be included regardless of disease or current prescription use, similar to the large-scale data mining approach used in Wang et al^18^. Additionally, we made use of a low parameter cosine only model because they are good for capturing internal circannual clock and effect of environmental changes (daylight, temperature, rainfall) and have one peak and one trough^48^. Sincos models would be useful when investigating specific tests more thoroughly where symmetric curves are not exhibited, or else multi-peak seasonality is expected. A low parameter approach was prioritized in this study as it is less test-specific and is more appropriate when taking a first attempt at a big data approach to modelling seasonality. In future studies, it would be beneficial to postulate higher-parameter algorithms to classes of tests in order to capture more specific and disease focused trends^48^.

Future studies in this area will also examine biochemical seasonality hand in hand with disease, prescription and gene expression seasonality, allowing for more personalized mathematical models to be fit to a patient’s medical history. Further, it would be beneficial to make use of future genotyping information as it is known to influence an individual’s healthy RI values^49^. As new information such as these become more widely available, improvements to the proposed model can be built overtime to create a reusable, generalized model which can be specified to any population regardless of deviations in hemisphere, dietary shifts, and weather patterns.

## Data Availability

The Danish National Patient Registry (DNPR) and laboratory data used can only be accessed by researchers authorized by the Danish health authorities. Approvals are listed in the manuscript.

## Abbreviations

BCC: B-Data Clinical Chemistry Laboratory System
CRP: c-reactive protein
CVD: Coronary Vascular Disease
DNK: code for Denmark
DL: Deep Learning
DNPR: Danish National Patient Registry
EHR: Electronic Health Record
FDR: false discovery rate
HDL Cholesterol: High Density Lipoprotein Cholesterol
ICD: International Statistical Classification of Diseases
IFCC: International Federation of Clinical Chemistry
LABKA: The Clinical Laboratory Information System
LOINC: Logical Observation Identifiers Names and Codes
ML: Machine Learning
NPU: Nomenclature, Properties and Units
NLS: non-linear least-squares
RI: Reference Interval
PSA: Prostate Specific Antigen
TSH: Thyroid Stimulating Hormone.

## Data Access Approval

This study has been approved by The Danish Data Protection Agency (ref: 514-0255/18-3000, 514-0254/18-3000, SUND-2016-50), The Danish Health Data Authority (ref: FSEID-00003724 and FSEID-00003092) and The Danish Patient Safety Authority (3-3013-1731/1/). The study has been approved as a registry study where patient consent is not needed in Denmark.

