## Supplemental table 1, figures 1+2 for "Population-wide analysis of laboratory tests to assess seasonal variation and the relevance of temporal reference interval modification"

#### Supplemental Material

**Supplemental Table 1: Overview of all parameter fits for amplitude and offset values.** Calculated as described in the methods, sex and age corrected fits are listed under "Overall Fits", sex specific (age corrected) parameter fits are listed under "Sex Fits", and mortality fits (sex and age corrected) are listed under "Mortality Fits." Fitted parameter values are bolded when the p_val for the fit is below 0.05 (FDR corrected). NA values are listed then there were not enough tests to fulfil threshold requirements described in methods, or else there was no data available, such as for sex-specific tests.

| TEST NAME | Overall Fits | | Sex Fits | | | | Mortality Fits | | | |
|---|---|---|---|---|---|---|---|---|---|---|
| | | | Male | | Female | | Alive | | Dead | |
| | Amplitude | Offset | Amplitude | Offset | Amplitude | Offset | Amplitude | Offset | Amplitude | Offset |
| ALANINE TRANSAMINASE (ALAT) - P | **0.02** | 2.45 | **0.02** | 4.28 | **0.02** | 0.84 | **0.02** | 2.75 | **0.02** | 5.88 |
| ALBUMIN - P | 0.01 | 6.95 | **0.01** | 6.67 | 0.01 | 7.10 | 0.01 | 5.28 | **0.01** | 4.41 |
| ALBUMIN - U | **0.02** | 3.57 | **0.02** | 5.60 | **0.03** | 2.47 | **0.02** | 3.27 | #N/A | #N/A |
| ALBUMIN / CREATININE; RATIO - U | **-0.04** | **11.44** | **-0.03** | **11.76** | **-0.04** | **11.89** | **-0.04** | **11.42** | #N/A | #N/A |
| ALKALINE PHOSPHATASE - P | **0.01** | **8.53** | **0.01** | **9.35** | **0.01** | **7.76** | **0.01** | **7.63** | **-0.03** | **52.00** |
| ALPHA-1 GLOBULIN - P | 0.02 | 5.91 | #N/A | #N/A | #N/A | #N/A | 0.01 | 6.51 | #N/A | #N/A |
| ALPHA-2 GLOBULIN - P | **0.01** | 5.81 | #N/A | #N/A | #N/A | #N/A | 0.01 | 7.99 | #N/A | #N/A |
| AMYLASE - P | 0.00 | 0.00 | **0.00** | 0.00 | 0.00 | 0.00 | **0.01** | 0.00 | -0.01 | **50.81** |
| AMYLASE PANCREAS TYPE - P | **0.01** | 6.52 | **0.01** | 6.63 | **0.01** | 3.93 | **0.01** | **6.77** | 0.02 | 7.62 |
| ANDROSTENEDIONE - P | -0.01 | 6.70 | #N/A | #N/A | #N/A | #N/A | -0.01 | 6.76 | #N/A | #N/A |
| ANION GAP (INCL. K +) - P | **0.01** | **10.94** | **0.02** | **11.03** | **0.01** | 8.37 | **0.01** | **10.76** | 0.01 | 8.15 |
| ANTIMULLERIAN HORMONE - P | **-0.08** | **40.41** | #N/A | #N/A | **-0.08** | **40.41** | **-0.08** | **40.42** | #N/A | #N/A |
| ANTITHROMBIN - P | **-0.01** | 2.12 | **-0.01** | 0.00 | -0.01 | 4.98 | 0.00 | 1.60 | #N/A | #N/A |
| ASPARTATE TRANSAMINASE [ASAT] - P | **0.02** | **19.49** | **0.02** | **19.63** | **0.02** | **18.42** | **0.02** | **19.48** | **0.05** | **26.17** |
| ATYPICAL CELLS - B | **0.04** | **9.17** | **0.04** | **8.30** | **0.04** | **8.58** | **0.04** | **8.94** | **0.03** | 7.27 |
| BASE EXCESS - Ecv | **0.01** | 8.63 | **0.01** | 6.31 | **0.03** | **10.03** | **0.02** | **10.72** | 0.04 | 0.00 |
| BASE EXCESS - P | 0.01 | 11.05 | **-0.05** | **41.91** | 0.01 | 5.09 | 0.02 | 15.85 | #N/A | #N/A |
| BASOPHILS - B | **-0.04** | **13.34** | **-0.03** | **14.32** | **-0.04** | **12.25** | **-0.03** | **13.91** | **0.04** | **41.81** |
| BETA-GLOBULIN - P | **0.02** | 0.00 | #N/A | #N/A | #N/A | #N/A | **0.02** | 0.00 | #N/A | #N/A |
| BICARBONATE - P | **0.00** | 3.32 | **0.00** | 2.39 | **0.00** | 4.60 | **0.00** | 2.82 | **0.01** | 6.56 |
| BICARBONATE - P(kB) | **0.01** | 0.00 | #N/A | #N/A | #N/A | #N/A | **0.01** | 0.00 | #N/A | #N/A |
| BILIRUBIN - P | **-0.01** | **8.76** | **-0.01** | 9.27 | **-0.01** | **13.66** | **0.00** | 7.84 | **-0.03** | **8.92** |
| BILIRUBIN - P(neonatal) | **0.02** | 0.00 | #N/A | #N/A | #N/A | #N/A | **0.02** | 0.00 | #N/A | #N/A |
| C-PEPTIDE - P | **-0.03** | 6.90 | #N/A | #N/A | #N/A | #N/A | **-0.03** | 6.72 | #N/A | #N/A |
| C-REACTIVE PROTEIN (CRP) - P | **0.03** | 5.57 | **0.03** | 6.82 | **0.03** | 4.68 | **0.03** | 7.02 | 0.10 | **52.00** |
| CALCIUM (ALBUMIN-CORRECTED) - P | **0.00** | 11.49 | **-0.01** | **9.56** | **0.00** | 11.33 | **0.00** | 11.26 | #N/A | #N/A |
| CALCIUM - P | **0.00** | **42.47** | **0.00** | 0.00 | **0.00** | **16.07** | **0.00** | **43.43** | **0.00** | 0.65 |
| CALCIUM ION-FREE - P | **0.00** | **11.69** | **0.00** | **10.99** | **0.00** | **13.52** | **0.00** | **12.47** | **0.00** | **10.03** |

| | | | | | | | | | | |
|---|---|---|---|---|---|---|---|---|---|---|
| CALPROTECTIN - F | -0.03 | 7.50 | #N/A | #N/A | #N/A | #N/A | -0.03 | 7.53 | #N/A | #N/A |
| CANCER ANTIGEN 125 (CA125) - P | 0.01 | 21.51 | #N/A | #N/A | 0.01 | 21.51 | 0.01 | 20.79 | #N/A | #N/A |
| CANCER ANTIGEN 19-9 (CA19-9) - P | -0.04 | 14.12 | #N/A | #N/A | #N/A | #N/A | -0.04 | 12.85 | #N/A | #N/A |
| CARBAMIDE - P | **0.01** | **12.29** | **0.01** | **12.64** | **0.01** | **12.29** | **0.01** | **13.15** | **0.04** | **5.98** |
| CARBAMIDE - U | 0.02 | 7.76 | **0.02** | 12.96 | #N/A | #N/A | 0.01 | 9.67 | #N/A | #N/A |
| CARBON MONOXIDE HEMOGLOBIN - Hb(B) | **0.01** | **19.05** | **-0.01** | 0.00 | **0.01** | **18.97** | **0.01** | **19.17** | 0.01 | 14.73 |
| CARCINOEMBRYONIC ANTIGEN; AMOUNT (CEA) - P | -0.01 | 4.46 | **0.02** | **23.85** | 0.01 | 20.05 | 0.00 | 1.83 | #N/A | #N/A |
| CHAIN LAMBDA (IG) FRIT - P | -0.03 | 13.80 | #N/A | #N/A | #N/A | #N/A | -0.03 | 13.18 | #N/A | #N/A |
| CHLORIDE - P | 0.00 | 0.00 | **0.00** | 6.75 | 0.00 | 6.08 | 0.00 | 7.67 | **0.00** | 6.62 |
| CHOLESTEROL HDL - P | **0.00** | **12.77** | **-0.01** | **12.25** | 0.00 | 14.73 | **0.00** | **13.22** | #N/A | #N/A |
| CHOLESTEROL LDL - P | **0.01** | 4.73 | **0.01** | 4.03 | **0.01** | 4.49 | **0.01** | 4.97 | #N/A | #N/A |
| CHOLESTEROL LDL - P(fPt) | 0.01 | 0.00 | #N/A | #N/A | #N/A | #N/A | 0.01 | 0.00 | #N/A | #N/A |
| CHOLESTEROL TOTAL - P | **0.01** | 0.00 | **0.01** | 1.09 | **0.01** | 0.00 | **0.01** | 0.17 | #N/A | #N/A |
| CHOLESTEROL VLDL - P | 0.00 | 0.00 | **0.01** | 0.00 | 0.00 | 0.00 | 0.00 | 0.00 | #N/A | #N/A |
| CHORIONIC GONADOTROPIN BETA [HCG] - P | 0.00 | 0.00 | #N/A | #N/A | 0.00 | 6.64 | 0.00 | **42.93** | #N/A | #N/A |
| CICLOSPORIN - B | -0.02 | 7.96 | **-0.03** | 3.65 | #N/A | #N/A | -0.02 | 5.64 | #N/A | #N/A |
| CO2 TOTAL - P(vB) | **0.00** | 4.07 | **0.01** | 3.98 | **0.00** | 4.08 | **0.00** | 3.97 | 0.00 | 7.73 |
| COAGULATION FACTOR II + VII + X (INR) - P | **-0.01** | **10.53** | **-0.01** | **13.23** | **-0.01** | **7.49** | **-0.01** | **10.66** | -0.01 | 11.48 |
| COAGULATION, SURFACE-INDUCED (APTT) - P | **0.00** | 0.00 | **0.01** | 0.16 | **0.01** | 0.00 | **0.01** | 0.00 | **0.01** | 0.00 |
| CREATINE KINASE MB (CKMB) - P | 0.01 | 18.10 | **0.01** | 18.48 | 0.02 | 15.10 | 0.01 | **20.42** | #N/A | #N/A |
| CREATINE KINASE TOTAL - P | **-0.02** | 0.00 | **-0.02** | 0.00 | **-0.03** | 0.35 | **-0.02** | 0.22 | **0.09** | 2.05 |
| CREATININE - P | **0.01** | 0.00 | **0.01** | 0.00 | **0.01** | 0.00 | **0.01** | 0.00 | **0.02** | 4.07 |
| CREATININE - Pt(U) | 0.00 | 43.63 | **0.01** | 0.00 | #N/A | #N/A | -0.01 | 13.39 | #N/A | #N/A |
| CREATININE - U | **0.03** | **6.04** | **0.03** | **6.18** | **0.04** | **5.89** | **0.03** | **6.07** | #N/A | #N/A |
| CREATININE CLEARANCE - Nyre | -0.01 | 8.28 | **-0.01** | 12.97 | #N/A | #N/A | -0.01 | 10.83 | #N/A | #N/A |
| CYCLIC CITRULLINATED PEPTIDE ANTIBODY (CCP) - P | 0.01 | 52.00 | **0.03** | **52.00** | 0.02 | 52.00 | 0.01 | 52.00 | #N/A | #N/A |
| D-DIMER - P | 0.01 | 14.19 | **0.01** | 14.77 | 0.02 | 12.94 | 0.01 | 20.63 | -0.11 | **40.48** |
| DEAMIDATED GLIADIN PEPTIDE ANTIBODY - P | **-0.02** | 0.57 | #N/A | #N/A | 0.00 | 0.00 | **-0.01** | 0.40 | #N/A | #N/A |
| DEHYDROEPIANDROSTERONE SULFATE - P | -0.01 | 9.53 | #N/A | #N/A | #N/A | #N/A | -0.01 | 9.75 | #N/A | #N/A |
| DNA (DOUBLE STRANDED) ANTIBODY - P | **0.02** | **15.40** | **0.03** | **17.02** | -0.01 | 0.00 | **0.02** | **15.61** | #N/A | #N/A |
| EOSINOPHILS - B | **-0.03** | **12.10** | **-0.03** | **12.26** | **-0.04** | **11.41** | **-0.03** | **11.34** | **-0.04** | **8.89** |
| ERYTHROBLASTS - B | **-0.29** | **14.63** | **-0.28** | **15.02** | **-0.32** | **14.46** | **-0.30** | **14.41** | #N/A | #N/A |
| ERYTHROCYTES - B | **0.01** | 1.87 | **0.01** | 1.72 | **0.01** | 1.58 | **0.01** | 1.69 | **0.01** | 8.09 |
| ERYTHROCYTES, VOL.FR. (EVF) - B | **0.00** | 1.70 | **0.00** | 1.04 | **0.00** | 1.00 | **0.00** | 3.38 | **0.01** | **12.21** |
| ERYTROCYTVOL. AGENT [MCV] - B | 0.00 | 0.00 | **0.00** | 3.18 | 0.00 | 4.23 | 0.00 | 14.66 | **0.00** | 5.40 |
| ERYTROCYTVOL. REL. SPREADING - Ercs(B) | **-0.01** | 0.00 | **-0.01** | 0.00 | **0.01** | **21.24** | **-0.01** | 0.00 | #N/A | #N/A |
| ESTRADIOL - P | **-0.02** | 12.33 | #N/A | #N/A | 0.01 | **40.06** | **-0.02** | 12.20 | #N/A | #N/A |
| ETHANOL - P | -0.02 | 14.63 | **0.02** | **42.30** | #N/A | #N/A | 0.01 | 0.00 | #N/A | #N/A |
| FERRITIN - P | **0.02** | 0.00 | **0.02** | 0.00 | **0.02** | 0.00 | **0.02** | 0.00 | #N/A | #N/A |
| FETOPROTEIN - P | **0.02** | 7.15 | **0.02** | 7.34 | #N/A | #N/A | **0.02** | 7.24 | #N/A | #N/A |
| FIBRINOGEN - P | **0.02** | 6.60 | **0.02** | **9.10** | **0.02** | 4.55 | **0.02** | 6.51 | #N/A | #N/A |
| FOLATES - P | **-0.02** | 0.00 | **-0.01** | 0.08 | **-0.02** | 0.00 | **-0.02** | 0.00 | #N/A | #N/A |
| FOLLITROPIN - P | 0.00 | 0.00 | #N/A | #N/A | 0.01 | 3.26 | 0.00 | 0.00 | #N/A | #N/A |
| FREE T3 INDEX - | **0.02** | 0.00 | **0.01** | 0.00 | **0.01** | 0.00 | **0.02** | 0.00 | #N/A | #N/A |
| GAMMA GLOBULIN - P | 0.01 | 4.21 | #N/A | #N/A | #N/A | #N/A | **0.02** | 7.23 | #N/A | #N/A |
| GLOMERULAR FILTRATION (EGFR) - Nyre | **0.00** | **17.42** | **0.00** | **18.64** | **0.00** | **17.31** | **0.00** | **17.79** | **-0.02** | 3.83 |
| GLUCOSE (120 MIN) OGTT - P(kB) | **-0.01** | 0.00 | #N/A | #N/A | **-0.01** | 0.00 | **-0.01** | 0.00 | #N/A | #N/A |
| GLUCOSE (FASTING) - P(vB; fPt) | 0.00 | 0.00 | **0.00** | **42.54** | 0.00 | 10.97 | 0.00 | 0.00 | #N/A | #N/A |

| | | | | | | | | | | |
|---|---|---|---|---|---|---|---|---|---|---|
| GLUCOSE (FROM HBA1C) - P | 0.00 | 0.00 | **0.00** | 18.67 | 0.00 | 3.53 | 0.00 | 0.00 | #N/A | #N/A |
| GLUCOSE - P | 0.00 | 1.00 | **0.00** | 0.00 | 0.00 | 1.57 | 0.00 | 1.69 | **0.01** | **15.46** |
| GLUCOSE - P(aB) | **0.01** | **7.25** | **0.01** | 5.77 | **0.01** | 8.12 | **0.01** | 6.23 | 0.01 | 10.86 |
| GLUCOSE - P(kB) | 0.00 | 2.76 | **0.00** | 3.54 | 0.00 | 0.00 | **0.00** | 2.38 | **-0.02** | 5.71 |
| GLUTAMYL TRANSFERASE - P | 0.00 | 0.00 | **0.01** | 6.27 | **-0.01** | 10.35 | 0.00 | 0.00 | #N/A | #N/A |
| HAPTOGLOBIN - P | **0.02** | **6.63** | **0.01** | 9.90 | **0.02** | 5.54 | **0.01** | 6.13 | #N/A | #N/A |
| HEMOGLOBIN - B | **0.00** | 0.23 | **0.00** | 0.15 | **0.00** | 1.35 | **0.00** | 0.05 | **0.01** | **8.75** |
| HEMOGLOBIN - Ercs(B) | **0.00** | 3.90 | **0.00** | 2.20 | **0.00** | 4.20 | **0.00** | 4.66 | **0.00** | 0.00 |
| HEMOGLOBIN - Rtcs(B) | 0.00 | **15.09** | **0.00** | **13.61** | 0.00 | **42.64** | 0.00 | **41.70** | #N/A | #N/A |
| HEMOGLOBIN A1C (HBA1C) - Hb(B) | 0.00 | 0.00 | **0.00** | **19.03** | 0.00 | 5.70 | 0.00 | 0.00 | #N/A | #N/A |
| HEPATITIS B VIRUS S-ANTIBODY - P | -0.05 | 5.62 | **-0.07** | 4.30 | -0.06 | 6.26 | **-0.06** | 6.04 | #N/A | #N/A |
| HOMOCYSTEINE - P | **0.01** | 0.00 | #N/A | #N/A | **0.02** | **44.66** | **0.01** | 0.00 | #N/A | #N/A |
| HYDROGEN ION - P(aB) | 0.00 | 6.79 | **0.00** | 7.27 | 0.00 | 0.00 | **0.00** | 3.86 | 0.00 | 3.45 |
| HYDROGEN ION - P(kB) | **0.00** | **11.40** | #N/A | #N/A | #N/A | #N/A | **0.00** | **11.90** | #N/A | #N/A |
| HYDROGEN ION - P(vB) | **0.00** | 4.03 | #N/A | #N/A | #N/A | #N/A | **0.00** | 4.10 | #N/A | #N/A |
| HYDROXYPROGESTERONE - P | 0.00 | 10.71 | #N/A | #N/A | #N/A | #N/A | 0.00 | 11.15 | #N/A | #N/A |
| IMMUNOGLOBULIN E - P | **-0.06** | **13.04** | #N/A | #N/A | 0.02 | 0.00 | **-0.06** | **13.09** | #N/A | #N/A |
| INHALATIONSANTIGENPANEL IGE - P | **-0.02** | 1.00 | #N/A | #N/A | -0.01 | 12.14 | **-0.02** | 0.99 | #N/A | #N/A |
| IRON - P | 0.00 | 0.00 | **0.01** | 0.00 | 0.00 | 0.00 | 0.01 | 0.00 | #N/A | #N/A |
| KAPPA CHAIN (IG) - P | **0.06** | 1.69 | #N/A | #N/A | #N/A | #N/A | **0.06** | 1.30 | #N/A | #N/A |
| LACTATE - P | **-0.01** | **14.40** | **-0.02** | **15.10** | -0.01 | 12.58 | **-0.01** | **12.28** | 0.01 | 43.66 |
| LACTATE - P(aB) | **-0.01** | **14.37** | **0.01** | **41.94** | 0.01 | **41.79** | **0.01** | **42.24** | **-0.03** | **9.52** |
| LACTATE - P(vB) | **0.01** | 0.00 | **0.01** | 0.00 | 0.01 | 0.00 | **0.01** | 0.00 | #N/A | #N/A |
| LACTATE DEHYDROGENASE (LDH) - P | **-0.01** | 0.00 | **-0.01** | **48.38** | **-0.01** | 1.78 | **-0.01** | 0.00 | -0.01 | 7.88 |
| LARGE UNSTAINED CELLS - B | **0.05** | 4.60 | **0.05** | 4.88 | **0.05** | 4.39 | **0.05** | 4.80 | 0.02 | 2.02 |
| LEUKOCYTES - B | **0.01** | 2.06 | **0.01** | 2.05 | **0.01** | 0.40 | **0.01** | 0.20 | **0.01** | **16.54** |
| LEUKOCYTES - U | **-0.01** | 6.73 | **-0.01** | 4.75 | 0.00 | 0.00 | **-0.02** | 7.43 | #N/A | #N/A |
| LITHIUM - P | **-0.01** | **14.42** | #N/A | #N/A | #N/A | #N/A | **-0.01** | **14.90** | #N/A | #N/A |
| LUTROPIN - P | **0.01** | **44.09** | #N/A | #N/A | 0.01 | 0.00 | **0.01** | **44.02** | #N/A | #N/A |
| LYMPHOCYTES - B | 0.00 | 0.00 | **0.01** | 0.00 | **-0.01** | 12.32 | 0.00 | 0.00 | 0.00 | **42.88** |
| LYMPHOS. + MONO. + BLASTS - B | **0.03** | 1.70 | **0.04** | 4.78 | #N/A | #N/A | **0.03** | 2.64 | #N/A | #N/A |
| MAGNESIUM - P | 0.00 | 14.19 | **0.00** | 0.00 | 0.00 | 11.67 | 0.00 | 13.57 | 0.00 | 4.65 |
| METAMYELO. + MYELOMA. + PROMYELOCYTES - B | **0.05** | 0.00 | 0.05 | 0.00 | 0.04 | 0.00 | 0.03 | 0.00 | 0.03 | 27.03 |
| METHYLMALONATE - P | 0.00 | 0.00 | **0.01** | **42.03** | 0.01 | 1.75 | 0.00 | 42.63 | #N/A | #N/A |
| MONOCYTES - B | **0.00** | 1.29 | **0.00** | 1.45 | **0.00** | 2.32 | **0.00** | 2.58 | -0.01 | 2.19 |
| MYOGLOBIN - P | -0.02 | 13.46 | **0.01** | 3.57 | **-0.06** | **12.95** | -0.02 | 11.09 | #N/A | #N/A |
| NAKED CORES - B | 0.00 | **43.84** | **0.00** | 0.00 | #N/A | #N/A | 0.00 | 0.00 | #N/A | #N/A |
| NEUTROPHILS - B | **0.01** | 2.33 | **0.01** | 3.43 | **0.01** | 1.52 | **0.01** | 0.93 | **0.02** | **17.84** |
| O2 SAT. - Hb | **-0.02** | **13.94** | **-0.01** | **14.64** | **-0.04** | **14.98** | **-0.02** | **14.44** | #N/A | #N/A |
| OROSOMUCOID - P | **0.02** | **10.33** | #N/A | #N/A | #N/A | #N/A | **0.02** | **10.30** | #N/A | #N/A |
| OXYGEN (O2) - Hb(ab) | **0.00** | 10.27 | **0.00** | 8.03 | **0.00** | 9.45 | **0.00** | 7.27 | 0.00 | 0.00 |
| OXYGEN (O2) - Hb(kb) | **0.01** | **14.27** | #N/A | #N/A | #N/A | #N/A | **0.01** | **14.53** | #N/A | #N/A |
| OXYGEN (O2) - Hb(vB) | **-0.03** | 0.78 | #N/A | #N/A | #N/A | #N/A | **-0.03** | 1.32 | #N/A | #N/A |
| OXYHEMOGLOBIN - Hb(Fe;tot.;aB) | 0.00 | 6.06 | **0.00** | 9.57 | 0.00 | 7.00 | **0.00** | 3.52 | 0.00 | 0.00 |
| OXYHEMOGLOBIN - Hb(Fe;tot.;vB) | **-0.03** | 0.85 | #N/A | #N/A | #N/A | #N/A | **-0.03** | 1.39 | #N/A | #N/A |
| PAPP A - P | 0.01 | 11.42 | #N/A | #N/A | 0.01 | 11.42 | 0.01 | 11.42 | #N/A | #N/A |
| PARATHYRIN [PTH] - P | **0.03** | **12.19** | **0.04** | **12.42** | **0.02** | **11.90** | **0.03** | **12.39** | #N/A | #N/A |
| PCO2 - P | **0.00** | 1.77 | **0.00** | 0.00 | **0.01** | 4.79 | 0.00 | 1.47 | **0.01** | 5.45 |

| | | | | | | | | | | |
|---|---|---|---|---|---|---|---|---|---|---|
| PCO2 - P(aB) | **0.01** | **7.06** | **0.01** | **5.42** | **0.01** | **7.95** | **0.01** | **5.90** | **0.01** | **7.64** |
| PCO2 - P(kB) | **-0.02** | **15.49** | **0.01** | **42.82** | **-0.02** | **14.01** | **-0.02** | **15.46** | #N/A | #N/A |
| PCO2 - P(vB) | **0.01** | 4.60 | **0.01** | 3.69 | #N/A | #N/A | **0.01** | 4.08 | #N/A | #N/A |
| PHOSPHATE (P; INORGANIC) - P | 0.00 | **16.38** | **0.00** | 15.60 | **0.00** | **17.66** | 0.00 | **17.28** | **0.01** | 6.46 |
| PLATELETS - B | **0.01** | **4.82** | **0.01** | **4.93** | **0.01** | **4.65** | **0.01** | **4.66** | 0.01 | 13.57 |
| PO2 - P | 0.00 | 15.44 | **0.00** | 8.76 | **-0.01** | **13.92** | 0.00 | 0.00 | -0.01 | 8.01 |
| PO2 - P(aB) | **-0.01** | **10.11** | **0.00** | 8.10 | **-0.01** | **11.24** | **-0.01** | **9.53** | 0.00 | **42.79** |
| PO2 - P(kB) | **0.02** | **10.29** | **0.02** | **10.71** | **0.02** | **10.97** | **0.02** | **10.17** | #N/A | #N/A |
| PO2 - P(vB) | **-0.03** | 1.69 | #N/A | #N/A | #N/A | #N/A | **-0.04** | 2.27 | #N/A | #N/A |
| POTASSIUM - P | 0.00 | 12.27 | **0.00** | 3.60 | 0.00 | 0.00 | 0.00 | 7.85 | **0.00** | 5.92 |
| PRO-BRAIN NATRIURETIC PEPTIDE [PRO-BNP] - P | **-0.07** | **41.56** | **-0.07** | **42.50** | #N/A | #N/A | 0.05 | 14.21 | #N/A | #N/A |
| PROCALCITONIN - P | 0.04 | 8.96 | 0.07 | 0.20 | #N/A | #N/A | 0.03 | 18.57 | #N/A | #N/A |
| PROLACTIN - P | -0.01 | 4.56 | #N/A | #N/A | -0.01 | 3.60 | -0.01 | 4.55 | #N/A | #N/A |
| PROSTATE SPECIFIC ANTIGEN (BOUND) (PSA) - P | **0.04** | 6.71 | **0.04** | 6.71 | #N/A | #N/A | **0.04** | 6.54 | #N/A | #N/A |
| PROSTATE SPECIFIC ANTIGEN (PSA) - P | 0.00 | 15.76 | **0.00** | 15.76 | #N/A | #N/A | 0.00 | 14.95 | #N/A | #N/A |
| PROTEIN - P | **0.00** | 3.60 | **0.00** | 1.49 | **0.00** | 3.00 | **0.00** | 4.25 | #N/A | #N/A |
| PROTEIN - Pt(U) | **0.05** | 0.00 | 0.06 | 0.00 | #N/A | #N/A | 0.04 | 0.00 | #N/A | #N/A |
| PROTEIN - U | 0.01 | 0.00 | **0.01** | 2.08 | 0.01 | 0.00 | 0.01 | 0.00 | #N/A | #N/A |
| PSA-FREE / TOTAL (PSA) - P | -0.03 | 13.04 | **-0.03** | 13.04 | #N/A | #N/A | -0.03 | 12.75 | #N/A | #N/A |
| RETICULOCYTES - B | 0.01 | 0.00 | **0.01** | 0.00 | 0.01 | **41.63** | 0.01 | 0.00 | #N/A | #N/A |
| RETICULOCYTES - Erc | **-0.02** | **14.65** | **0.02** | **44.03** | **-0.02** | 10.54 | **0.02** | **42.29** | #N/A | #N/A |
| SEDIMENTATION REACTION - B | **0.04** | **13.22** | **-0.04** | **39.80** | **0.03** | **12.30** | **0.03** | **13.52** | #N/A | #N/A |
| SEXUAL HORMONE BINDING GLOBULIN (SHBG) - P | **0.04** | 0.00 | #N/A | #N/A | **0.05** | 0.00 | **0.04** | 0.00 | #N/A | #N/A |
| SODIUM - P | **0.00** | 2.61 | **0.00** | 1.60 | **0.00** | 1.83 | 0.00 | 1.91 | **0.00** | 1.22 |
| SODIUM - U | 0.00 | 1.61 | **0.00** | 0.00 | #N/A | #N/A | 0.00 | 7.35 | #N/A | #N/A |
| TACROLIMUS - B | 0.01 | 0.16 | **-0.01** | 0.00 | **0.03** | 3.20 | 0.01 | 0.82 | #N/A | #N/A |
| TESTOSTERONE - P | **-0.01** | 5.45 | **-0.01** | 9.63 | **-0.02** | 2.37 | **-0.01** | 5.24 | #N/A | #N/A |
| THYROID AUTOANTIBODIES (ANTI TPO) - P | **0.03** | **21.15** | #N/A | #N/A | **-0.03** | **47.85** | **0.02** | **21.48** | #N/A | #N/A |
| THYROID AUTOANTIBODIES (TRAB) - P | 0.00 | 3.22 | #N/A | #N/A | 0.00 | 3.22 | 0.00 | 3.22 | #N/A | #N/A |
| THYROTROPIN (TSH) - P | **0.03** | 1.59 | **0.03** | 1.42 | **0.03** | 1.56 | **0.03** | 1.65 | **0.03** | 0.00 |
| THYROXINE FREE [T4] - P | -0.01 | 0.00 | **0.01** | 20.23 | -0.01 | 0.00 | -0.01 | 0.00 | #N/A | #N/A |
| THYROXINE [T4] - P | 0.01 | 0.00 | **0.01** | 0.00 | 0.00 | 0.00 | 0.01 | 0.00 | #N/A | #N/A |
| TRANSFERRIN - P | 0.00 | 3.36 | **0.00** | 11.30 | **0.00** | 0.94 | 0.00 | 3.63 | #N/A | #N/A |
| TRANSFERRIN SATURATION - P | 0.01 | 0.00 | **0.00** | 0.00 | 0.01 | 0.00 | 0.01 | 0.00 | #N/A | #N/A |
| TRIGLYCERIDE - P | **0.02** | 0.00 | **0.03** | 0.00 | **0.01** | 0.00 | **0.02** | 0.00 | #N/A | #N/A |
| TRIGLYCERIDE - P(fPt) | **0.03** | 0.00 | **0.04** | 0.00 | **0.02** | 0.00 | **0.03** | 0.00 | #N/A | #N/A |
| TRIIODOTHYRONINE FREE [T3] - P | **-0.01** | 3.94 | **-0.01** | 4.25 | **-0.01** | 3.80 | **-0.01** | 3.78 | #N/A | #N/A |
| TRIIODOTHYRONINE REACTION - P | 0.00 | **43.62** | **0.00** | 0.55 | 0.01 | **42.61** | 0.00 | **43.70** | #N/A | #N/A |
| TRIIODOTHYRONINE [T3] - P | 0.00 | **42.60** | **0.01** | 0.94 | 0.00 | 12.81 | 0.00 | 0.00 | #N/A | #N/A |
| TROPONIN I - P | 0.01 | 9.89 | **0.02** | **14.11** | 0.01 | 9.32 | 0.00 | 11.53 | #N/A | #N/A |
| TROPONIN T - P | **0.03** | **12.87** | **0.03** | 13.64 | **0.05** | **14.02** | **0.04** | **14.15** | #N/A | #N/A |
| URATE - P | **-0.01** | 2.32 | **-0.01** | 0.39 | **-0.01** | 2.95 | **-0.01** | 2.40 | 0.01 | 18.41 |
| VITAMIN B12 - P | **0.04** | **3.08** | **0.04** | **3.15** | **0.04** | **2.98** | **0.04** | **3.07** | #N/A | #N/A |
| VITAMIN D(CALCIFEDIOL) - P | **-0.09** | **8.38** | **-0.11** | **8.18** | **-0.08** | **8.57** | **-0.09** | **8.36** | #N/A | #N/A |
| ZINC - P | 0.01 | 1.58 | **0.01** | 3.90 | 0.00 | 0.00 | **0.01** | 6.80 | 0.01 | 10.74 |

### Supplemental Figure 1

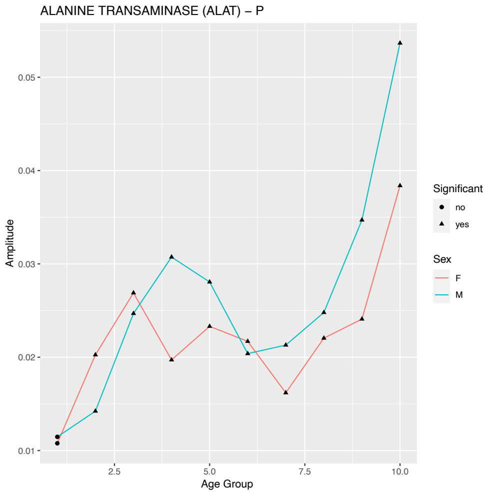


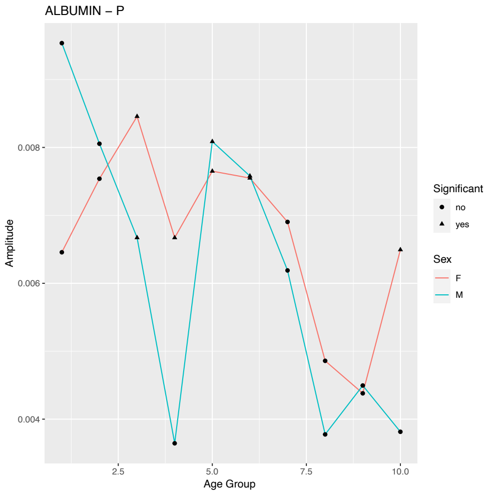


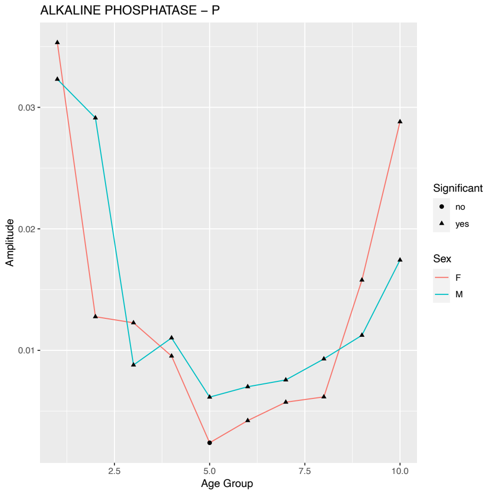


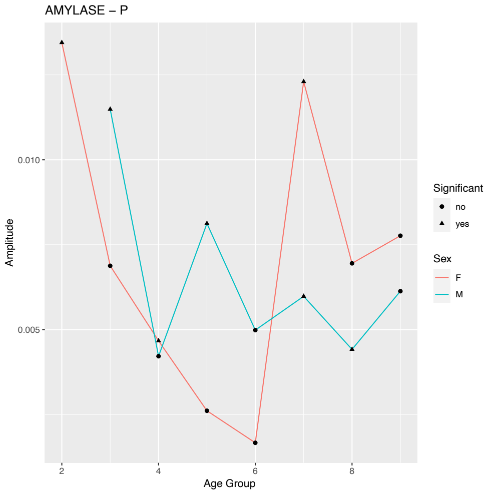


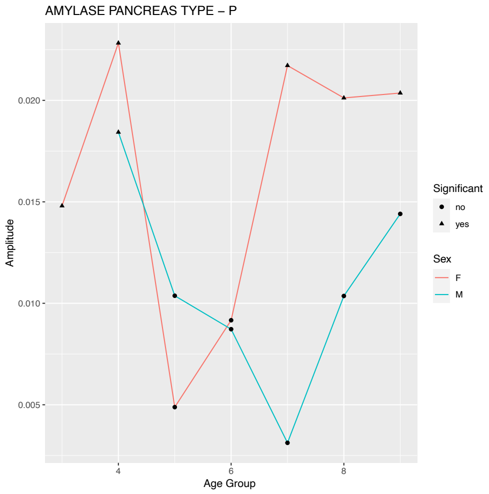


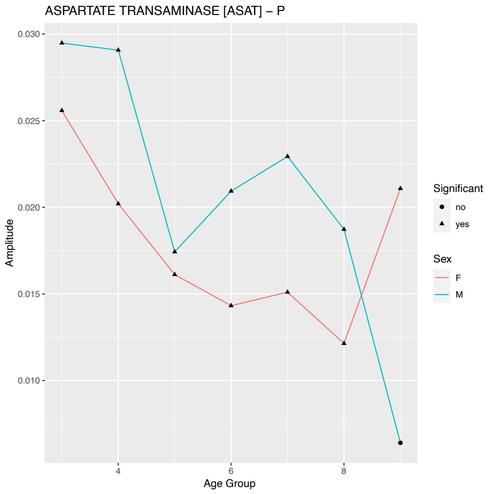

### Supplemental Figure 1

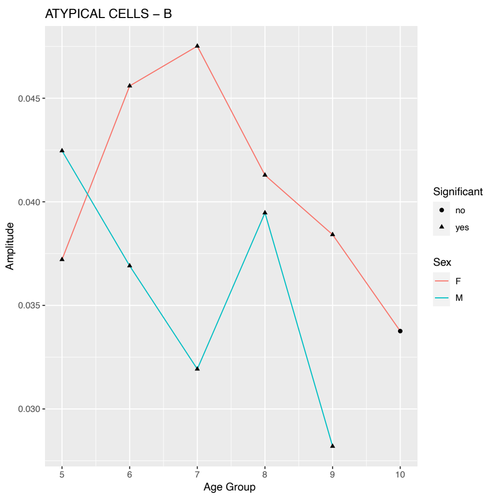


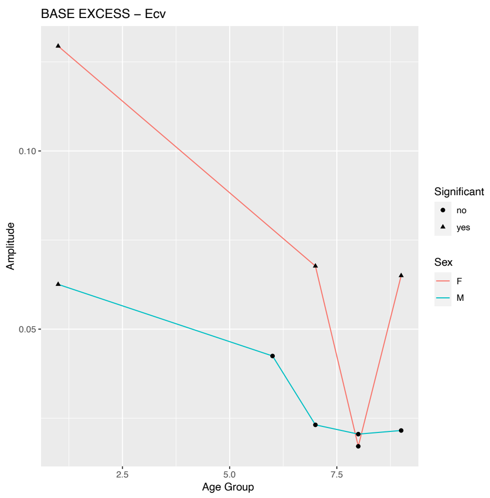


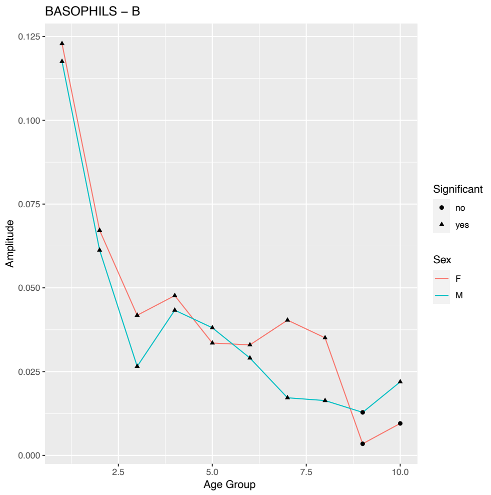


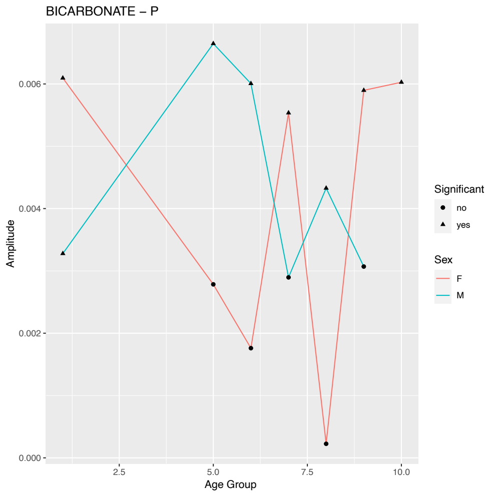


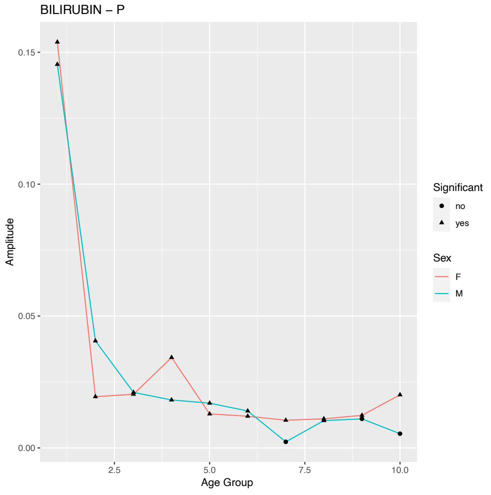


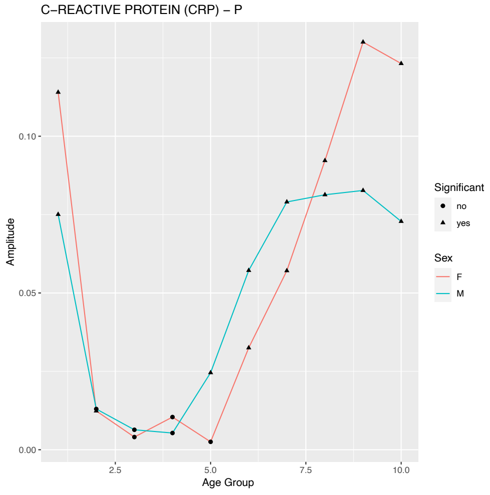

### Supplemental Figure 1

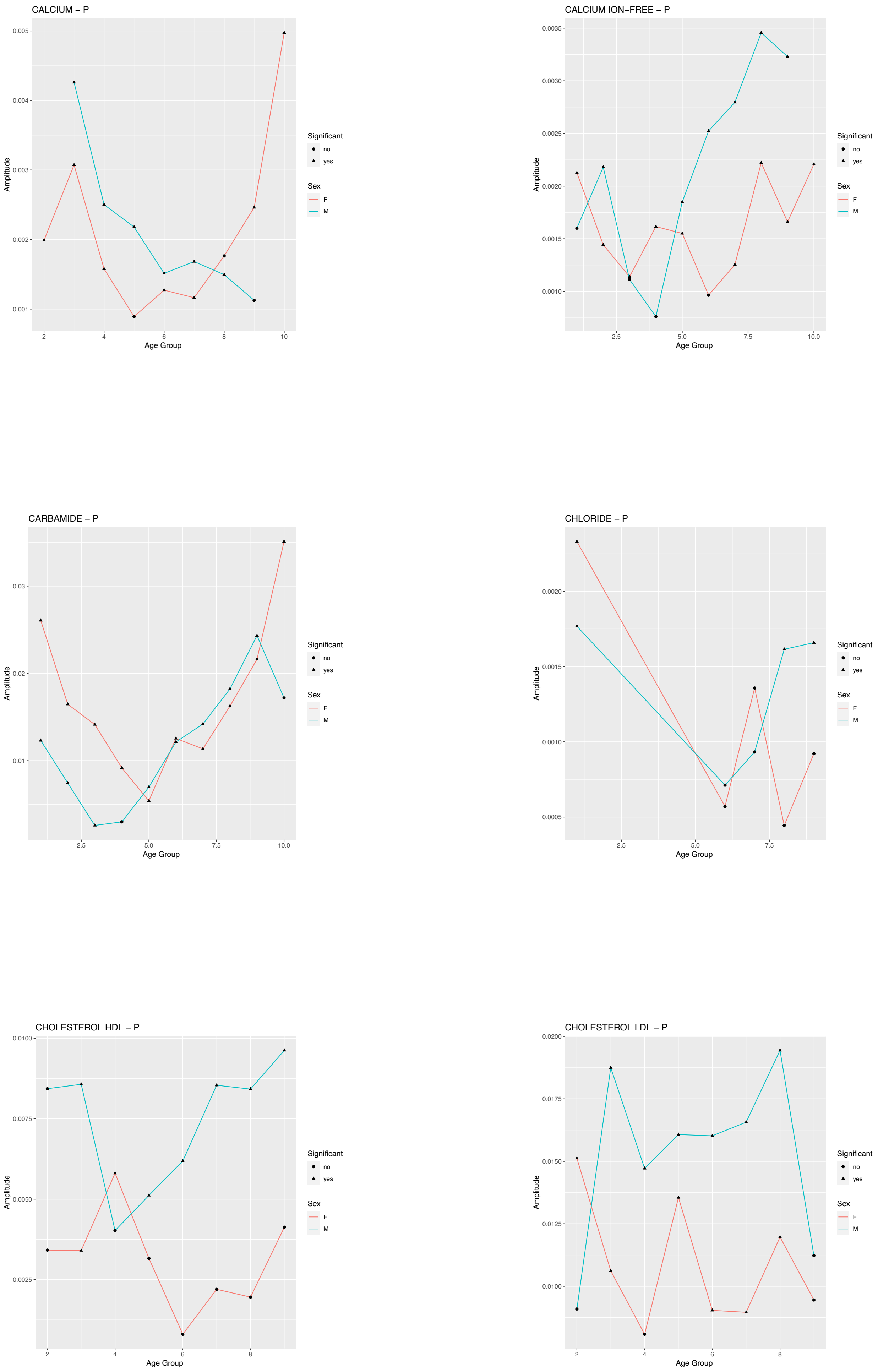

### Supplemental Figure 1

CHOLESTEROL TOTAL – P

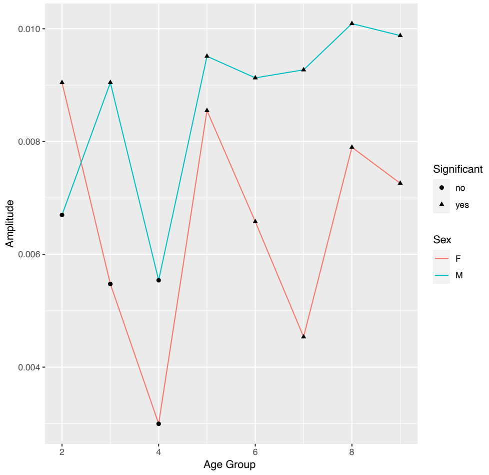


CHOLESTEROL VLDL – P

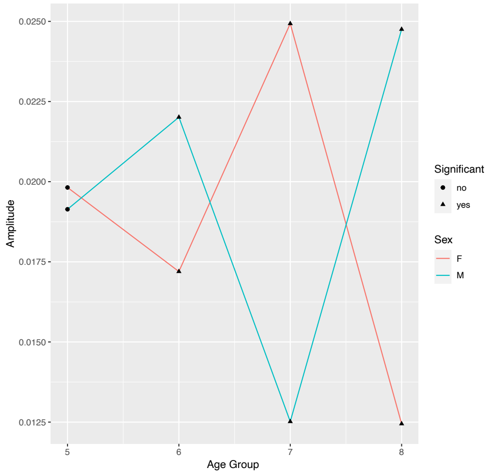


CO2 TOTAL – P(vB)

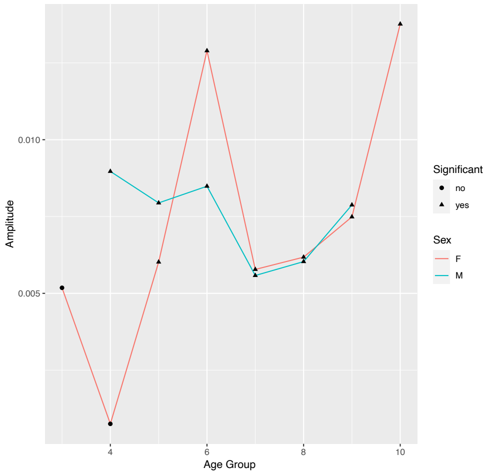


COAGULATION FACTOR II + VII + X (INR) – P

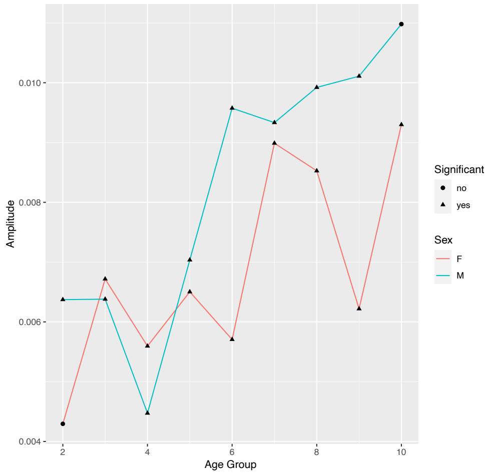


COAGULATION, SURFACE–INDUCED (APTT) – P

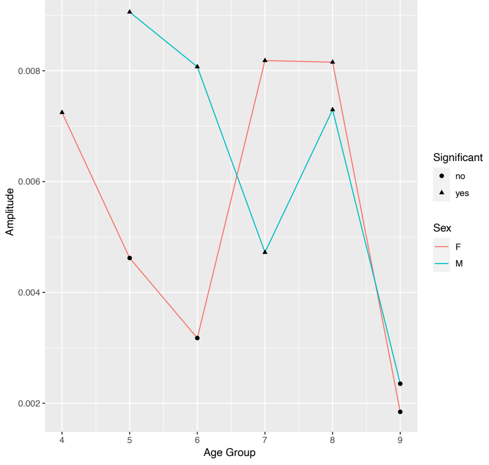


CREATINE KINASE TOTAL – P

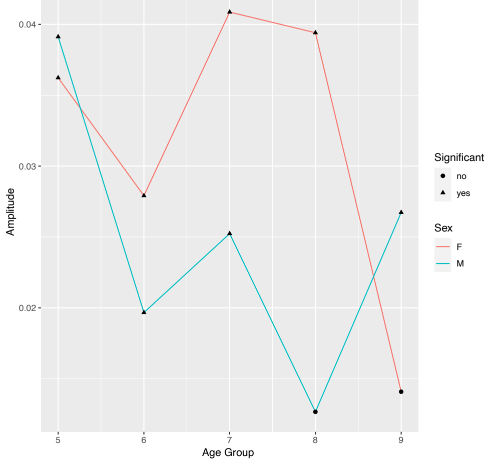

### Supplemental Figure 1

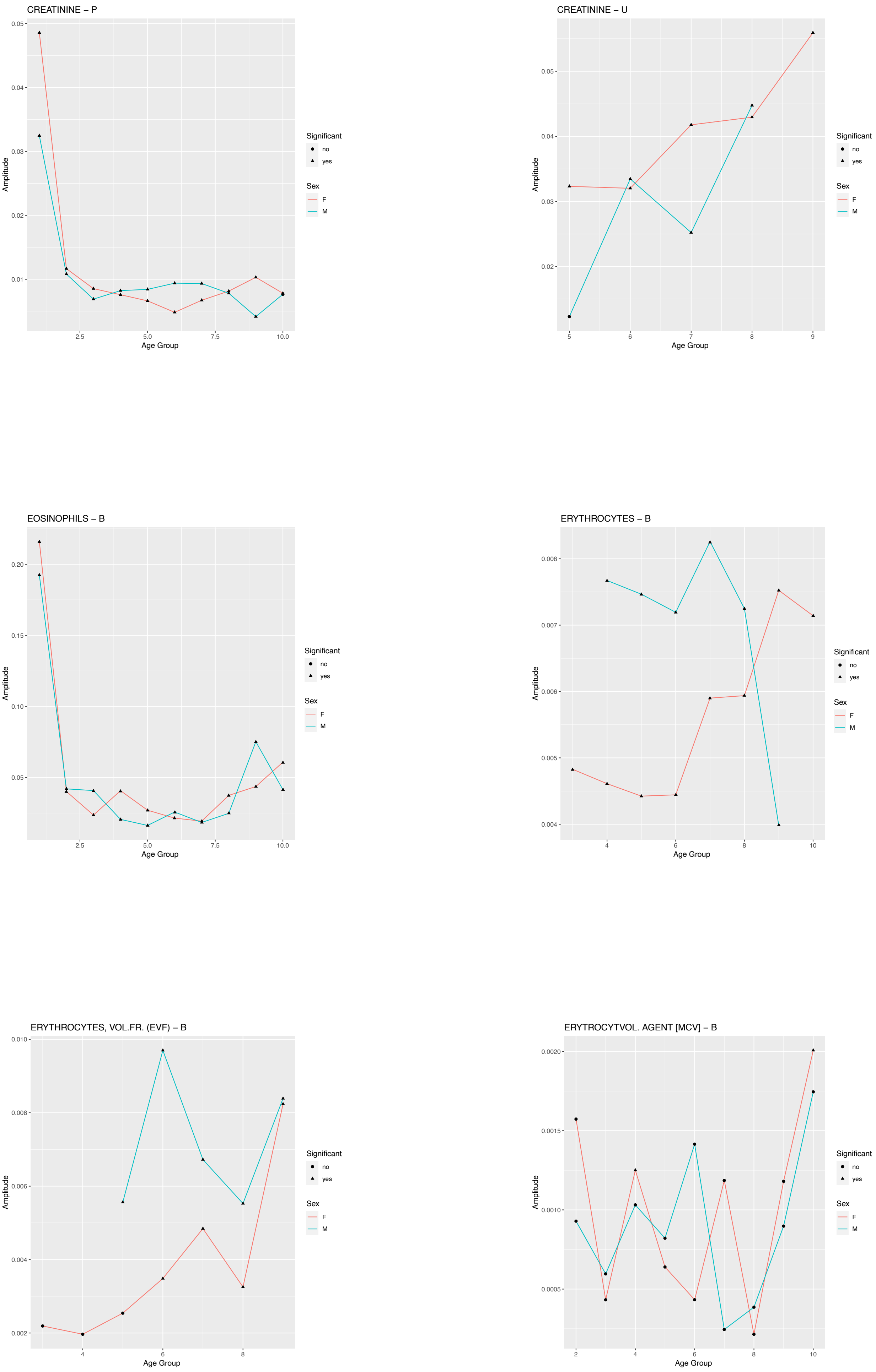

### Supplemental Figure 1

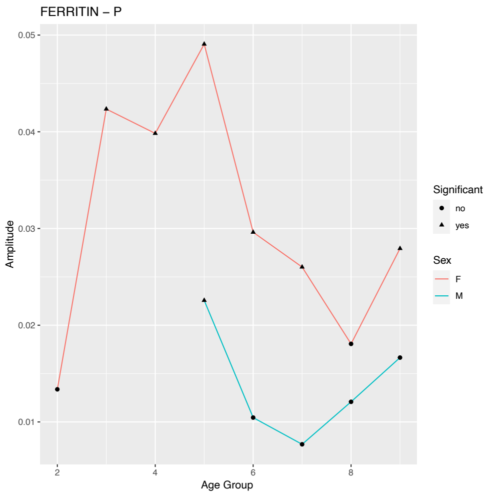


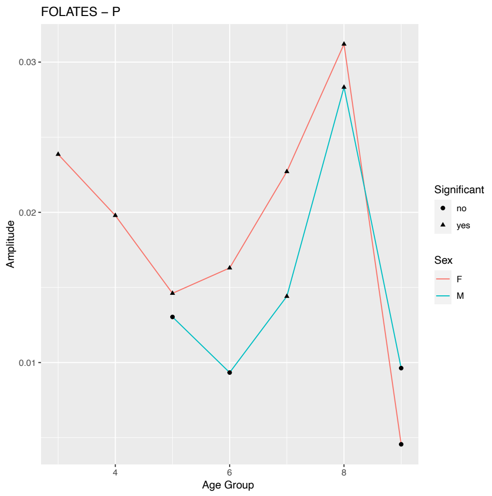


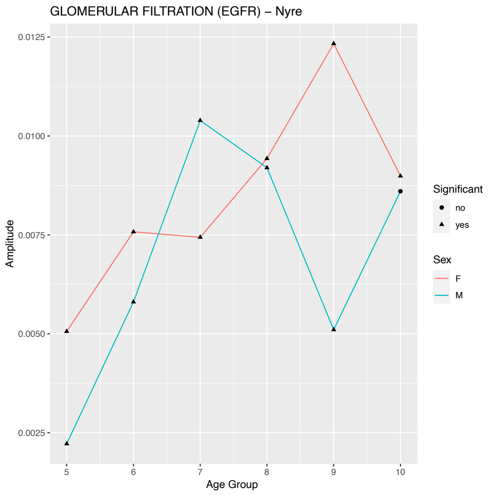


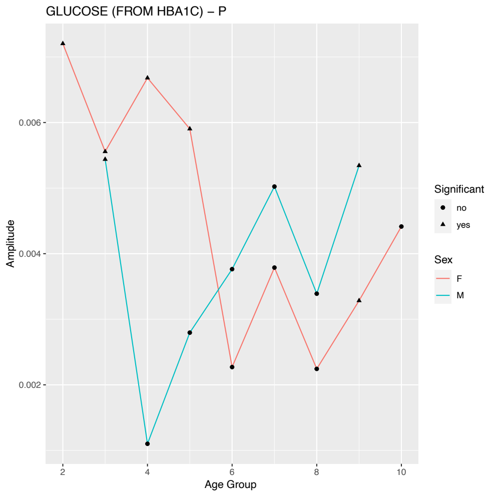


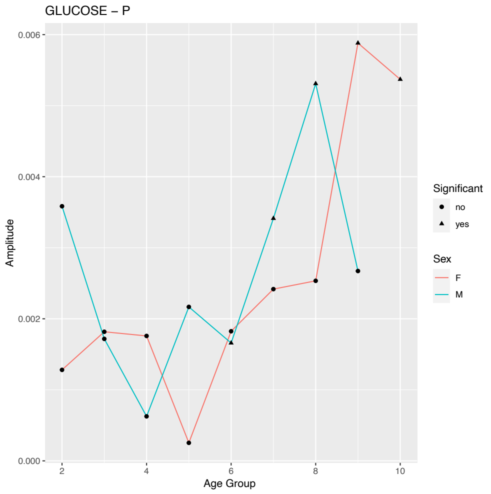


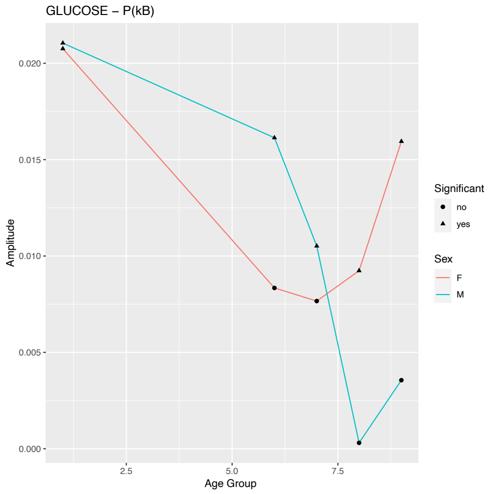

### Supplemental Figure 1

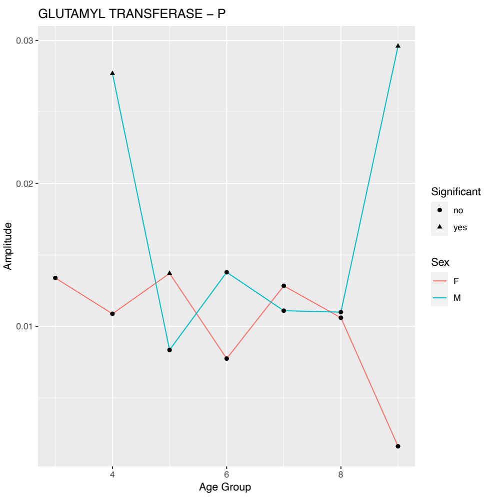


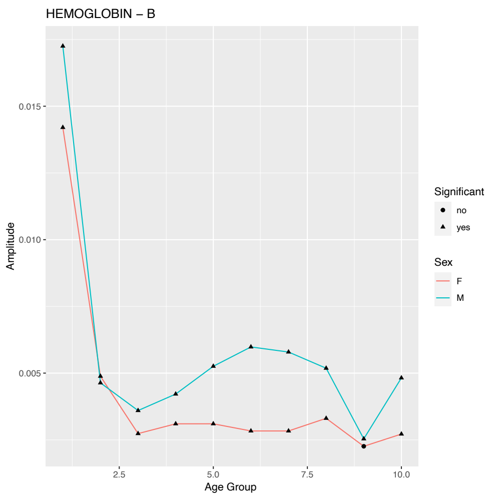


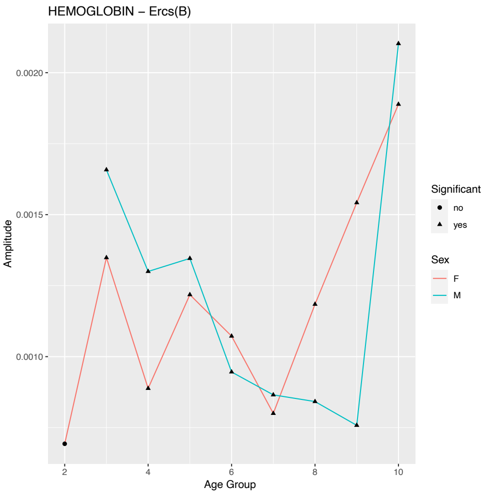


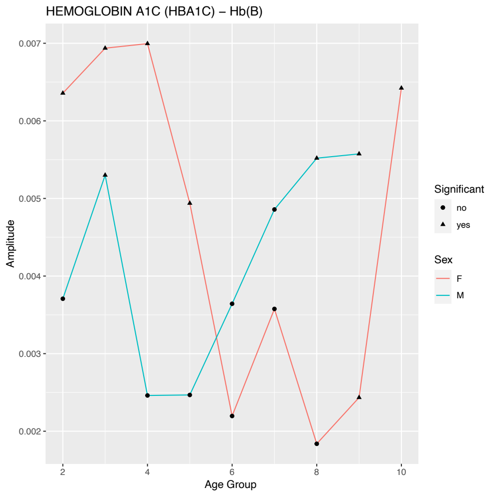


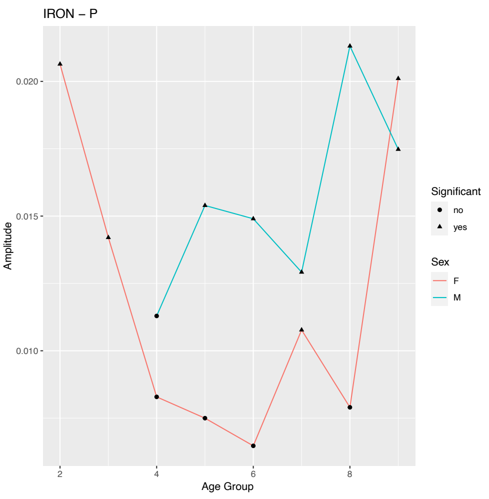


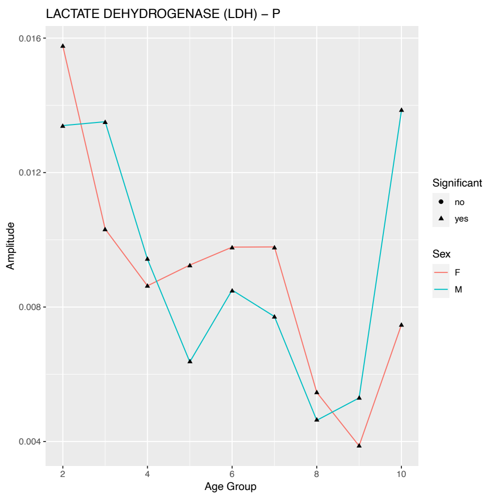

### Supplemental Figure 1

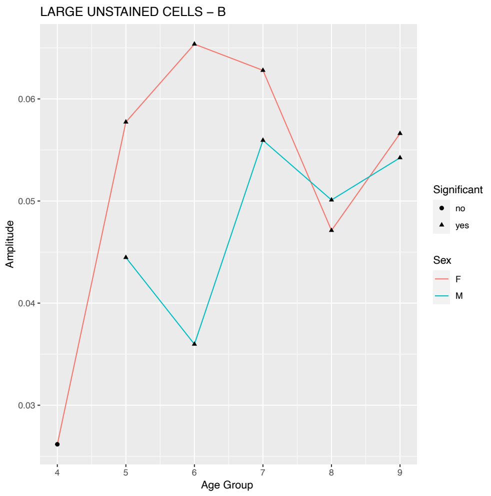


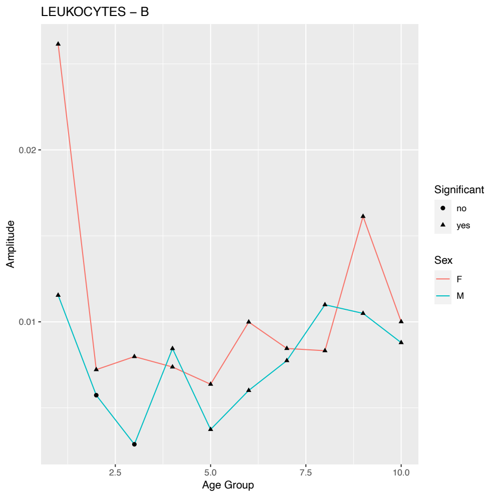


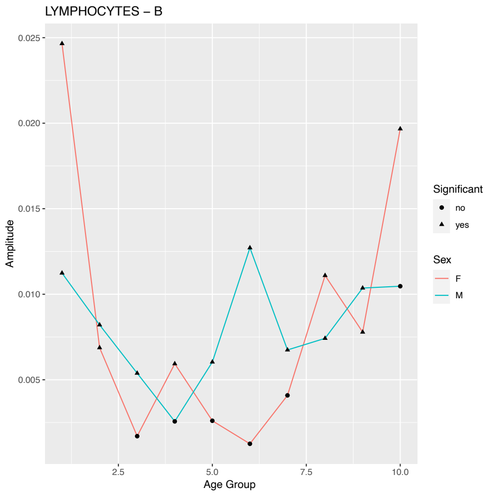


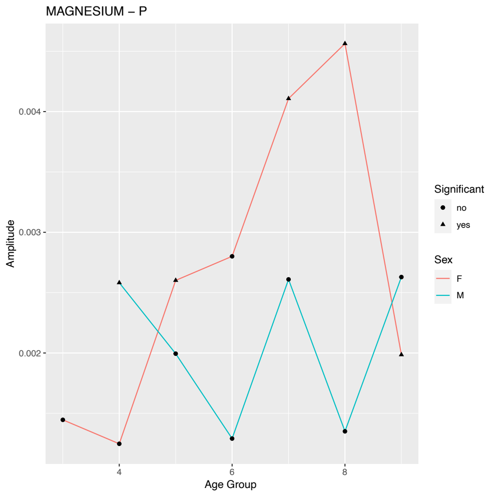


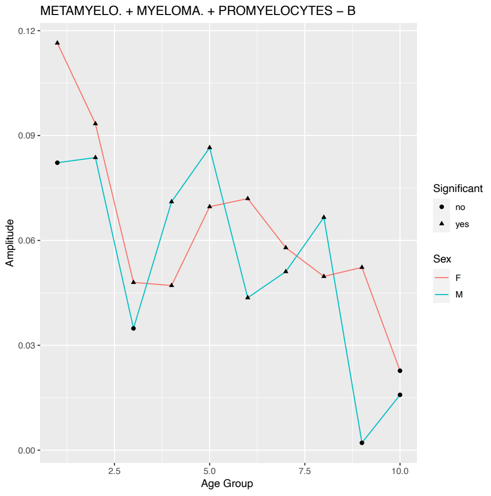


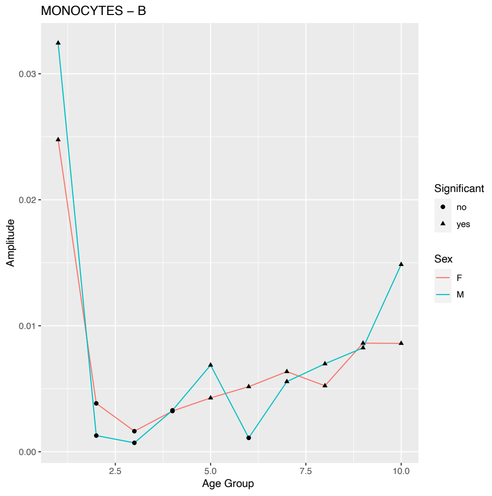

### Supplemental Figure 1

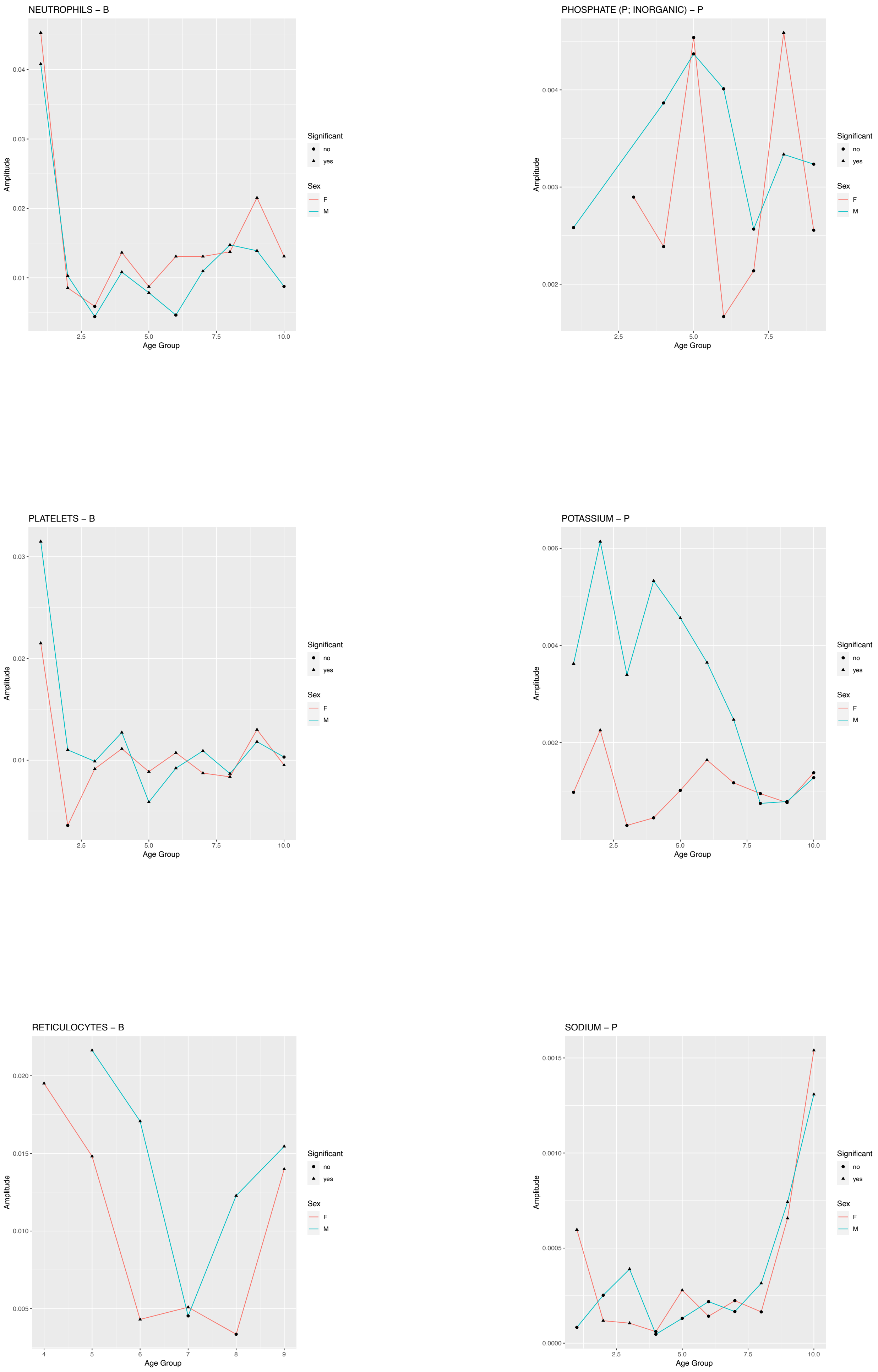

### Supplemental Figure 1

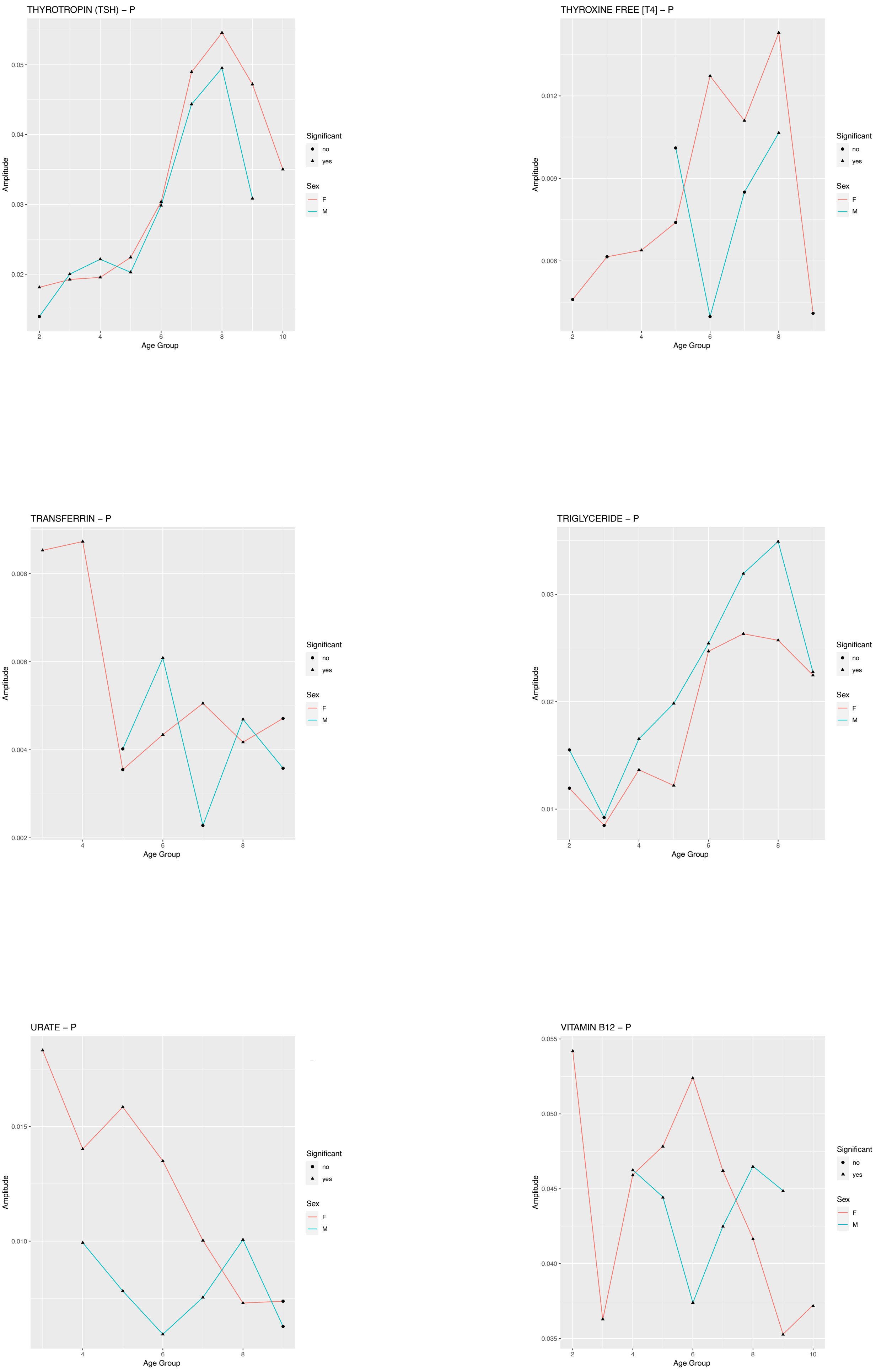

**Supplemental Figure 1**

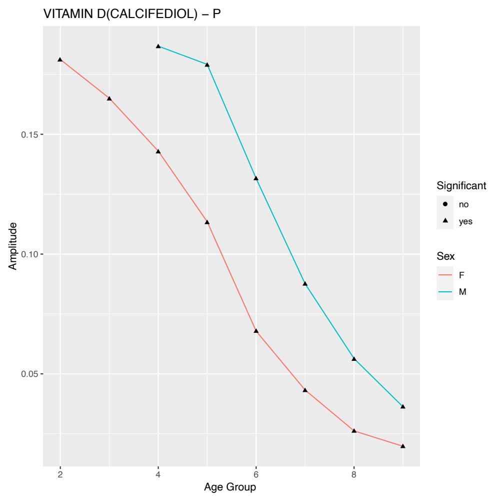


**Supplemental Figure 1: Overview of amplitude fits by age and sex group.** The figure shows the absolute value of the fitted amplitude variable per unique test, age group, and sex, alphabetically. Closed triangles indicate significant fits while closed circles indicate non-significant amplitude fits, using FDR corrected p-values and a threshold for significance of <0.05. Only tests with at least 4 different age group data points and both sexes represented were included in this figure in order to properly discern sex and age differences.

#### Supplemental Figure 2

ALANINE TRANSAMINASE (ALAT) – P

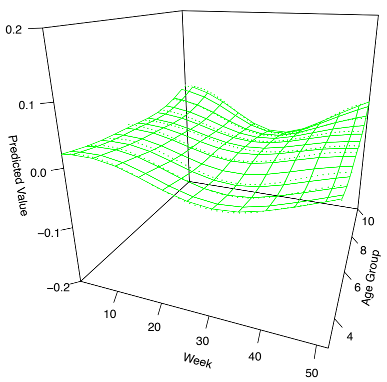


ALBUMIN – P

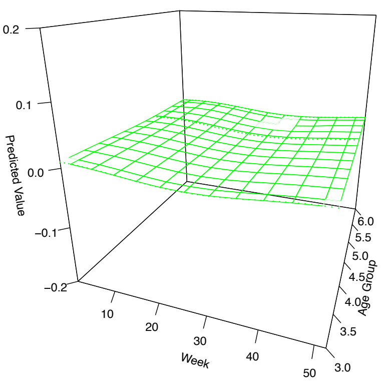


ALBUMIN – U

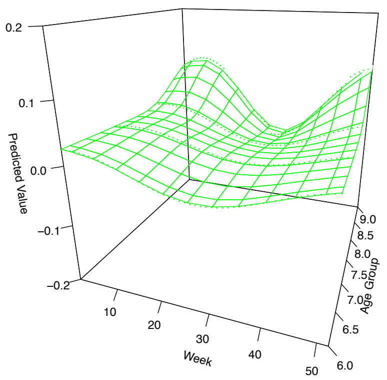


ALBUMIN / CREATININE; RATIO – U

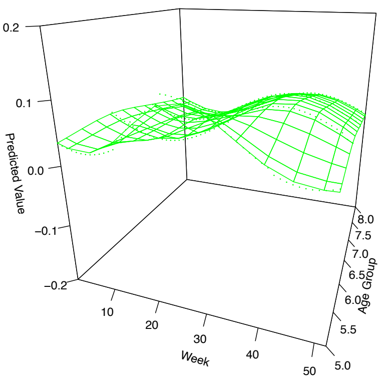


ALKALINE PHOSPHATASE – P

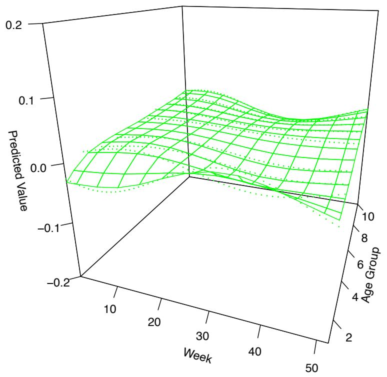


AMYLASE – P

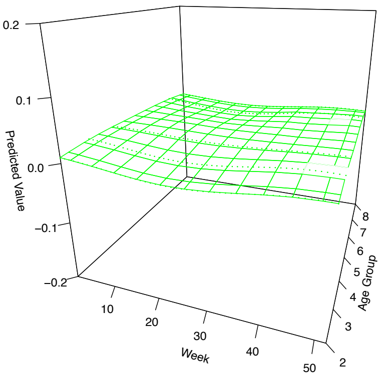

#### Supplemental Figure 2

**AMYLASE PANCREAS TYPE – P**

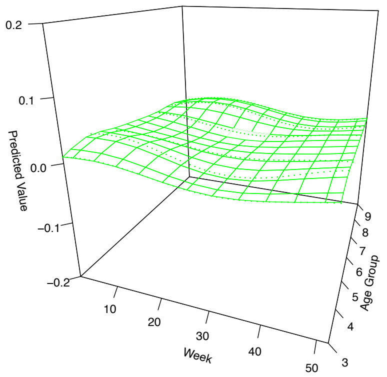


**ASPARTATE TRANSAMINASE [ASAT] – P**

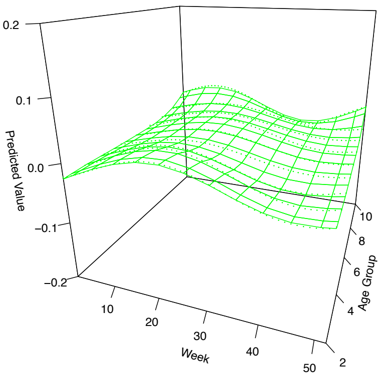


**ATYPICAL CELLS – B**

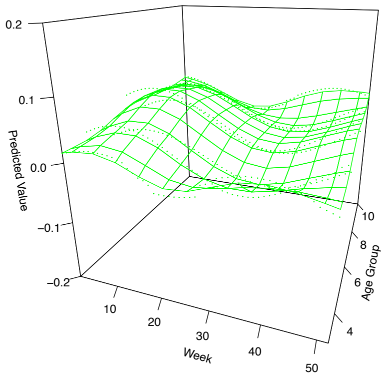


**BASE EXCESS – Ecv**

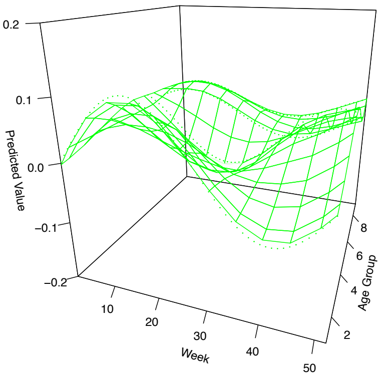


**BASOPHILS – B**

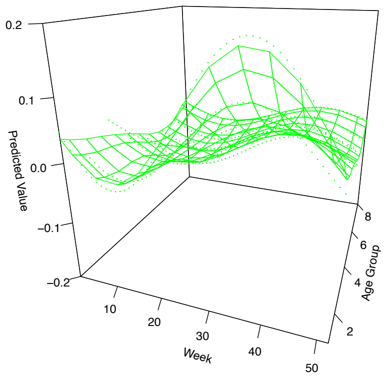


**BICARBONATE – P**

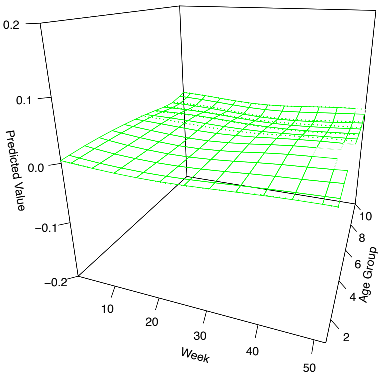

#### Supplemental Figure 2

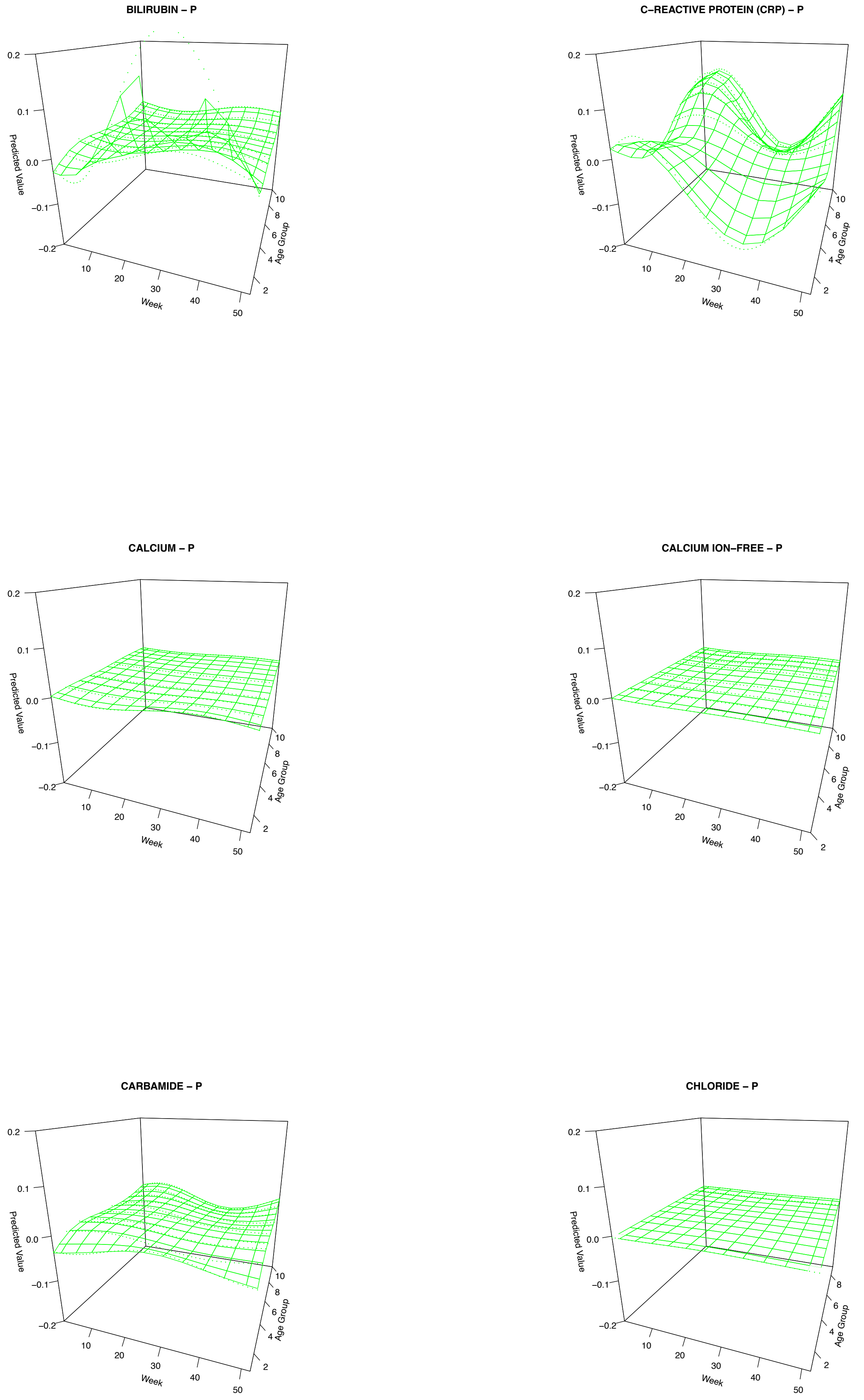

#### Supplemental Figure 2

CHOLESTEROL HDL – P

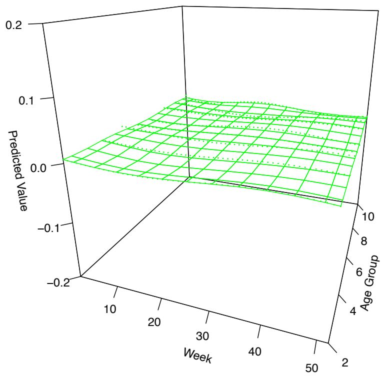


CHOLESTEROL LDL – P

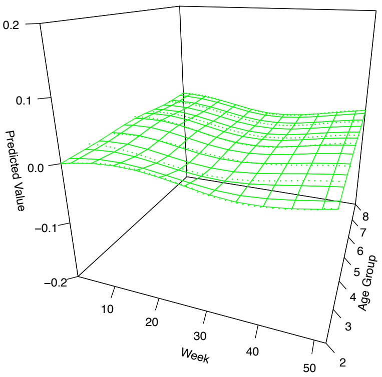


CHOLESTEROL TOTAL – P

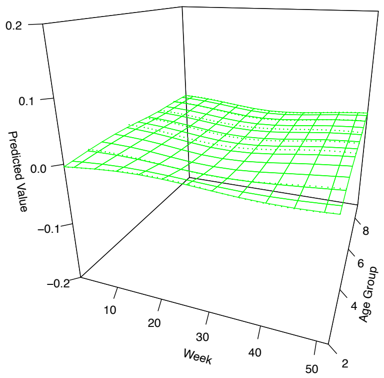


CHOLESTEROL VLDL – P

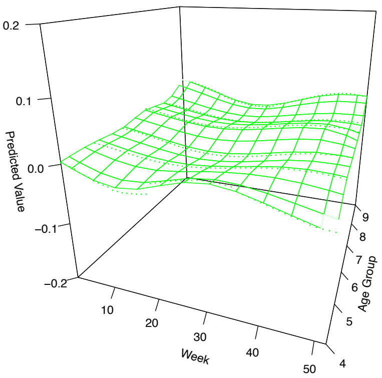


CO2 TOTAL – P(vB)

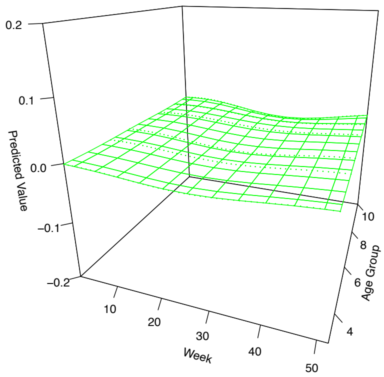


COAGULATION FACTOR II + VII + X (INR) – P

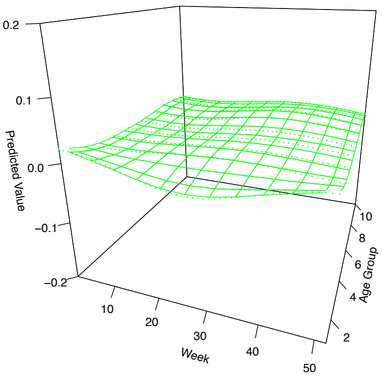

### Supplemental Figure 2

**COAGULATION, SURFACE–INDUCED (APTT) – P**

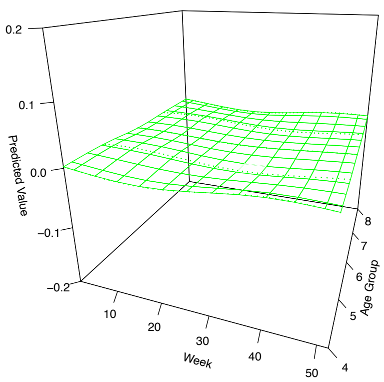


**CREATINE KINASE MB (CKMB) – P**

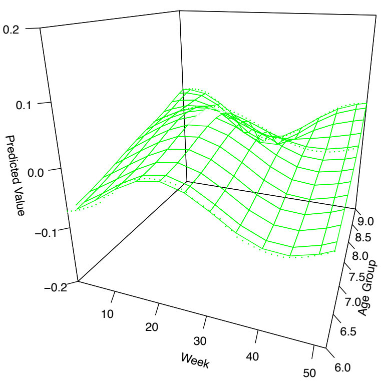


**CREATINE KINASE TOTAL – P**

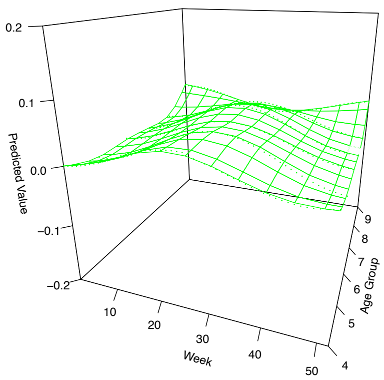


**CREATININE – P**

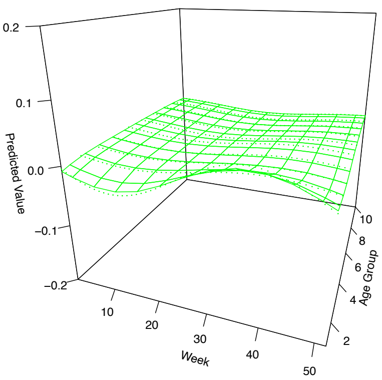


**CREATININE – U**

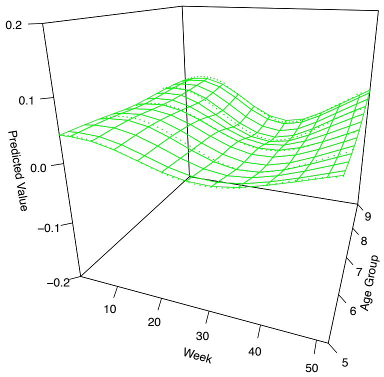


**D–DIMER – P**

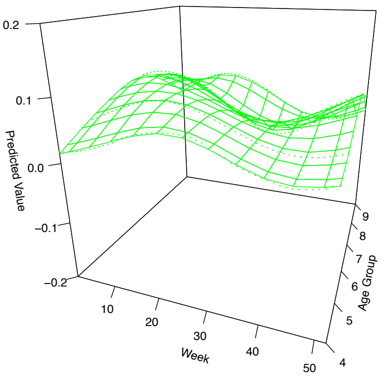

### Supplemental Figure 2

EOSINOPHILS – B

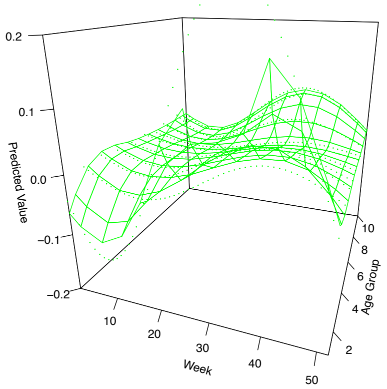


ERYTHROCYTES – B

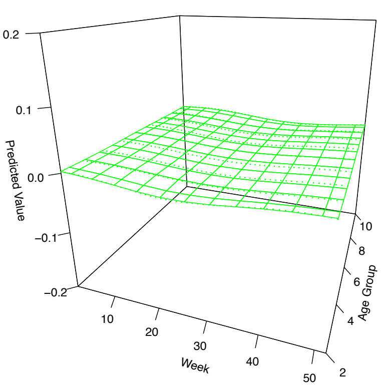


ERYTHROCYTES, VOL.FR. (EVF) – B

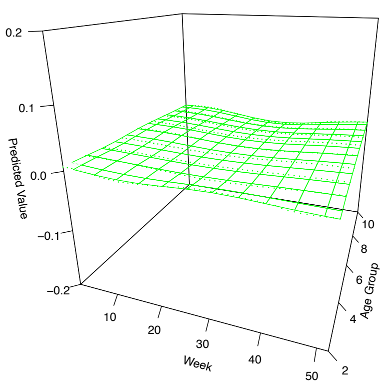


ERYTROCYTVOL. AGENT [MCV] – B

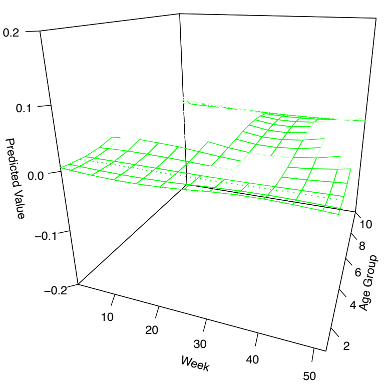


FERRITIN – P

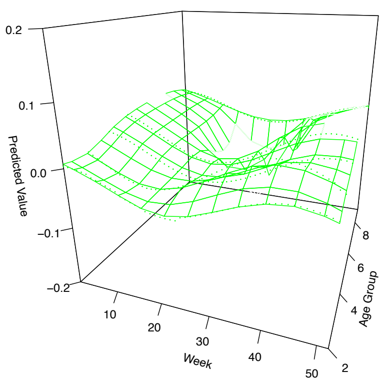


FOLATES – P

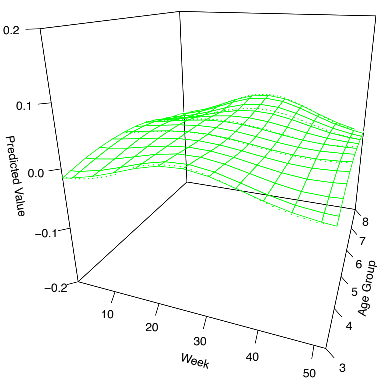

#### Supplemental Figure 2

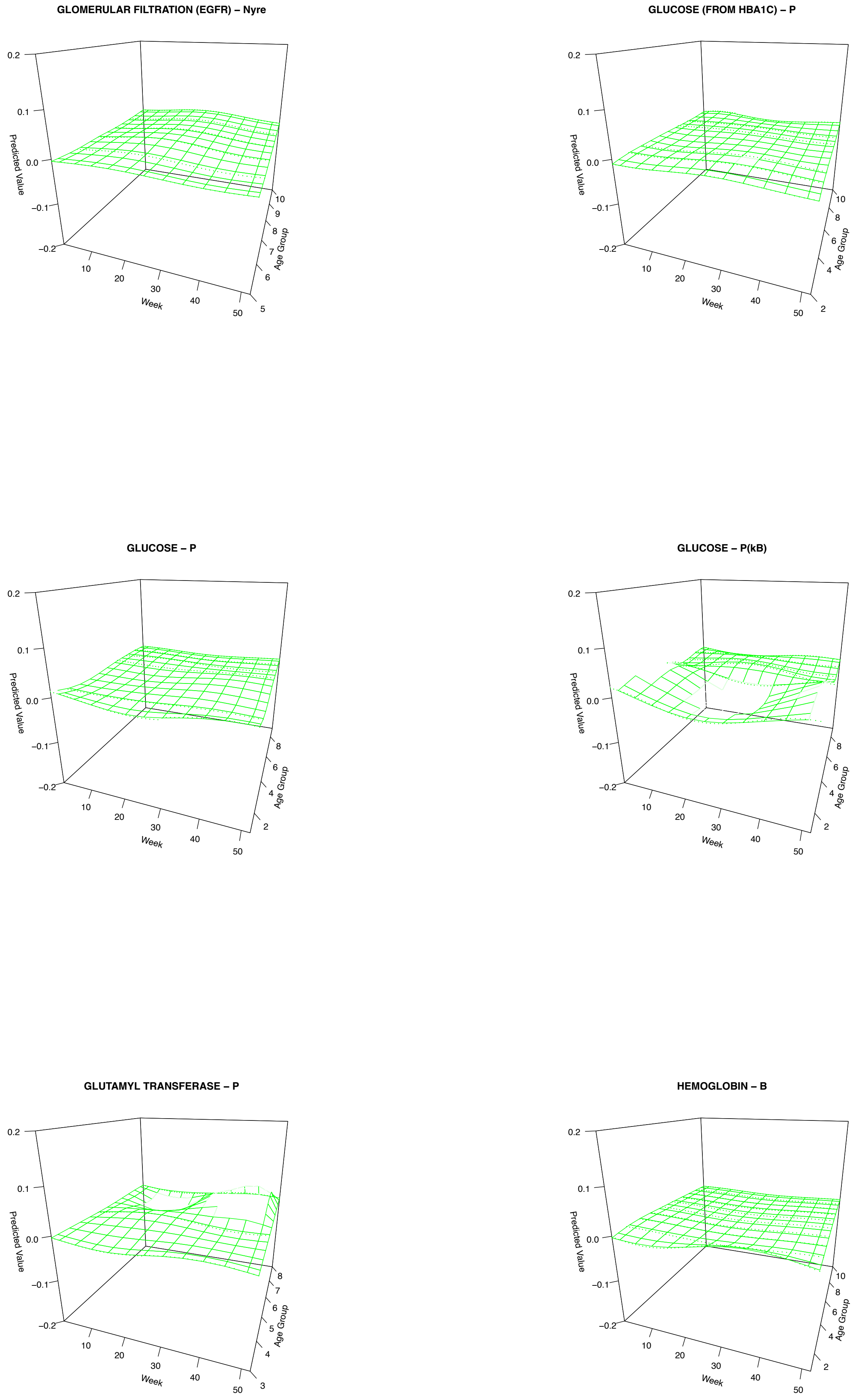

### Supplemental Figure 2

HEMOGLOBIN – Ercs(B)

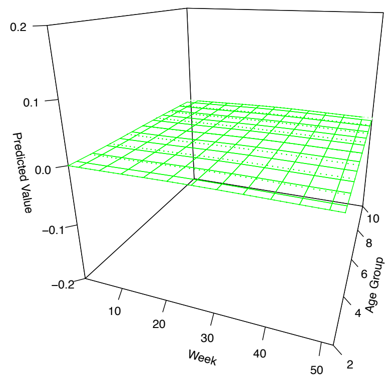


HEMOGLOBIN A1C (HBA1C) – Hb(B)

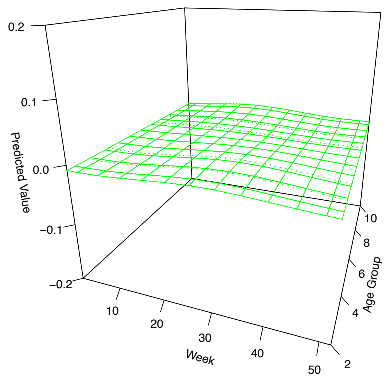


IRON – P

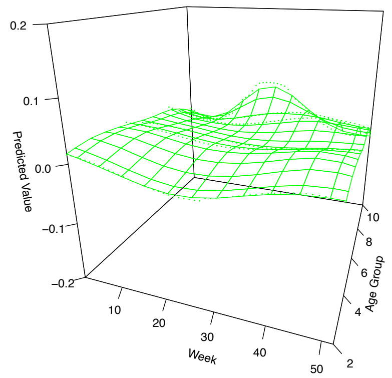


LACTATE DEHYDROGENASE (LDH) – P

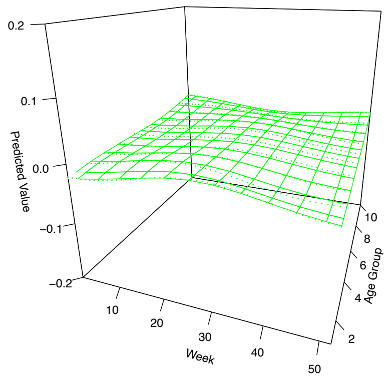


LARGE UNSTAINED CELLS – B

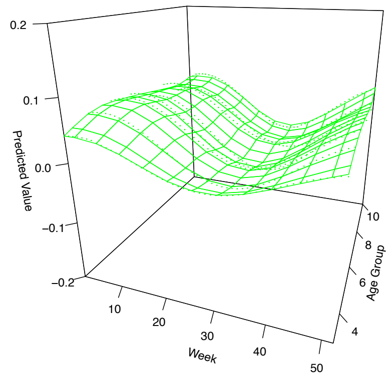


LEUKOCYTES – B

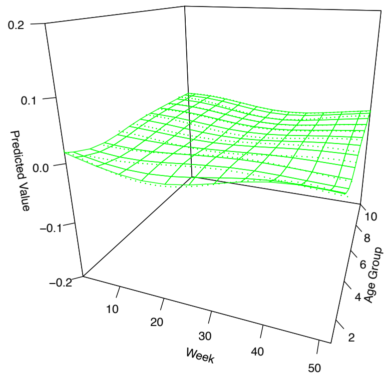

#### Supplemental Figure 2

**LYMPHOCYTES – B**

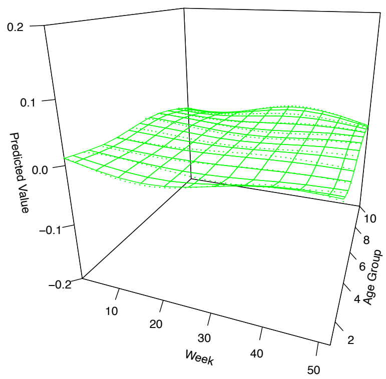


**METAMYELO. + MYELOMA. + PROMYELOCYTES – B**

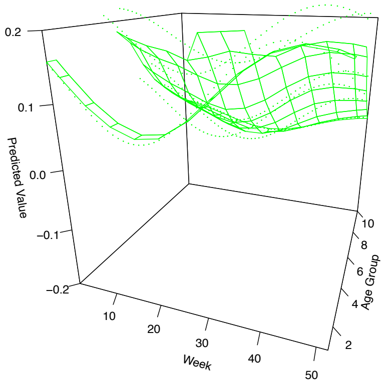


**MONOCYTES – B**

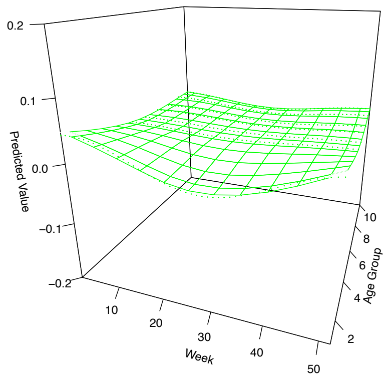


**NEUTROPHILS – B**

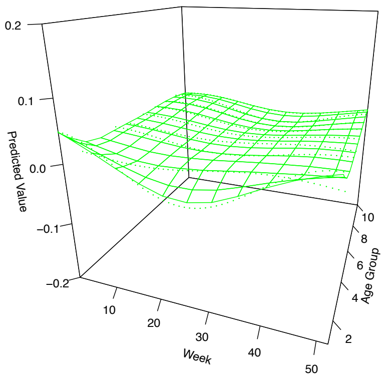


**PARATHYRIN [PTH] – P**

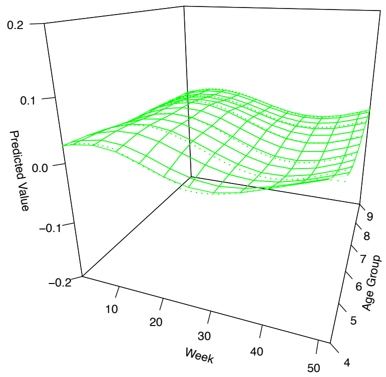


**PCO2 – P**

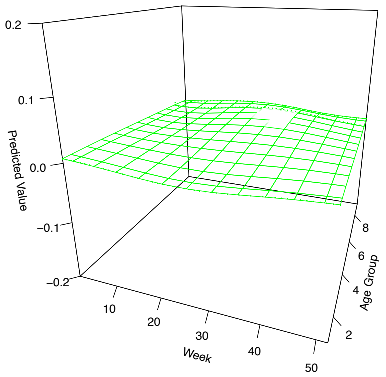

### Supplemental Figure 2

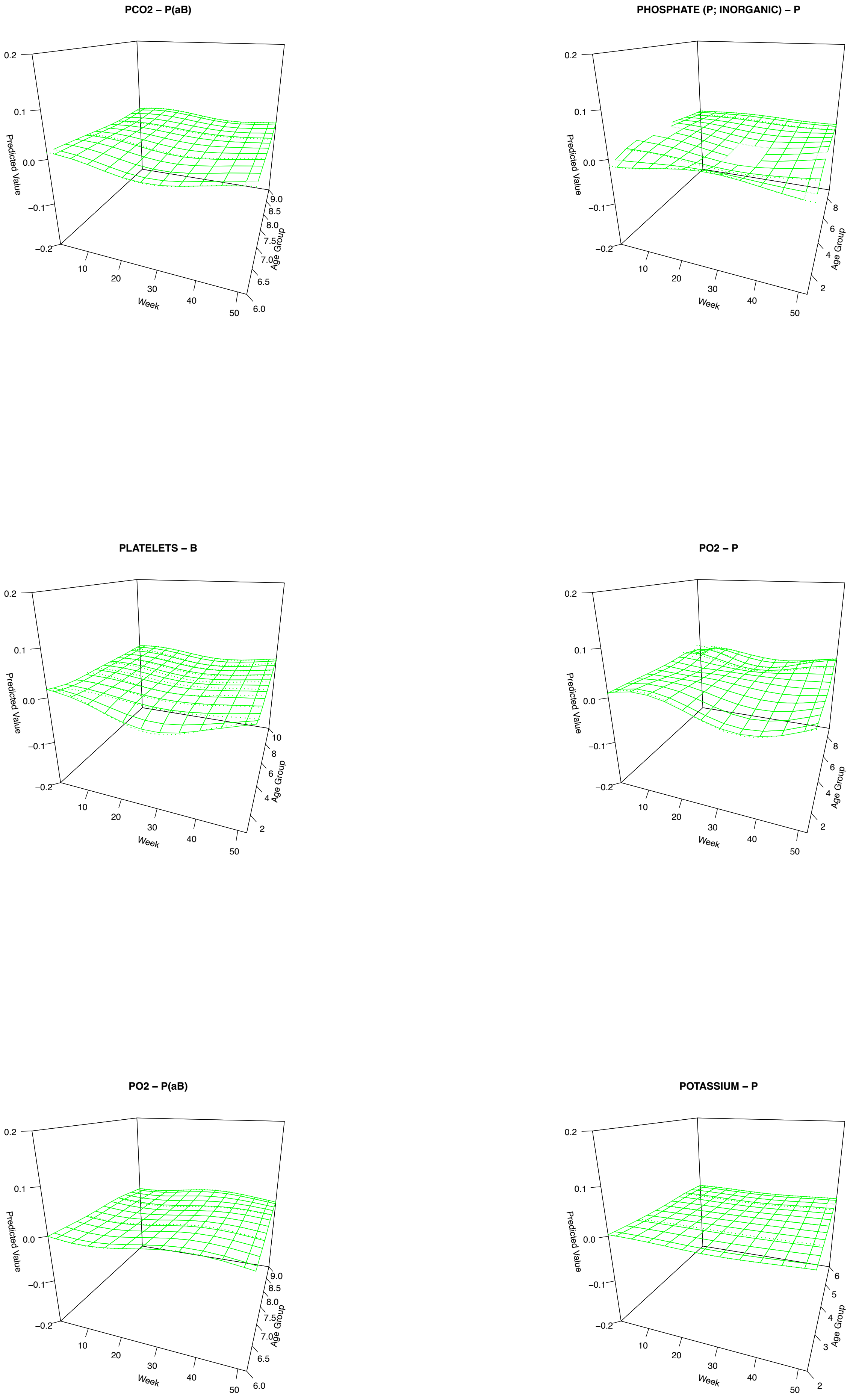

### Supplemental Figure 2

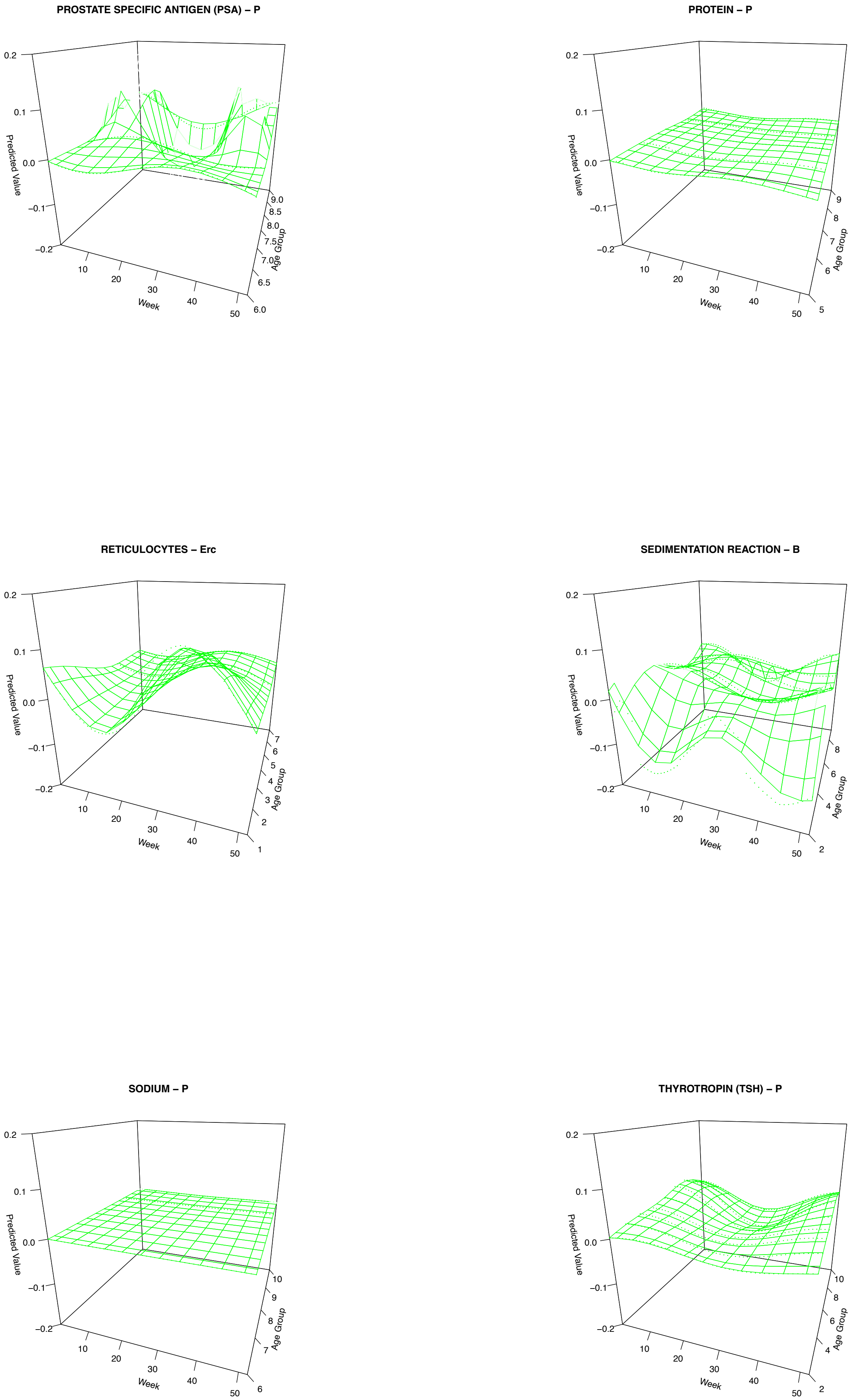

#### Supplemental Figure 2

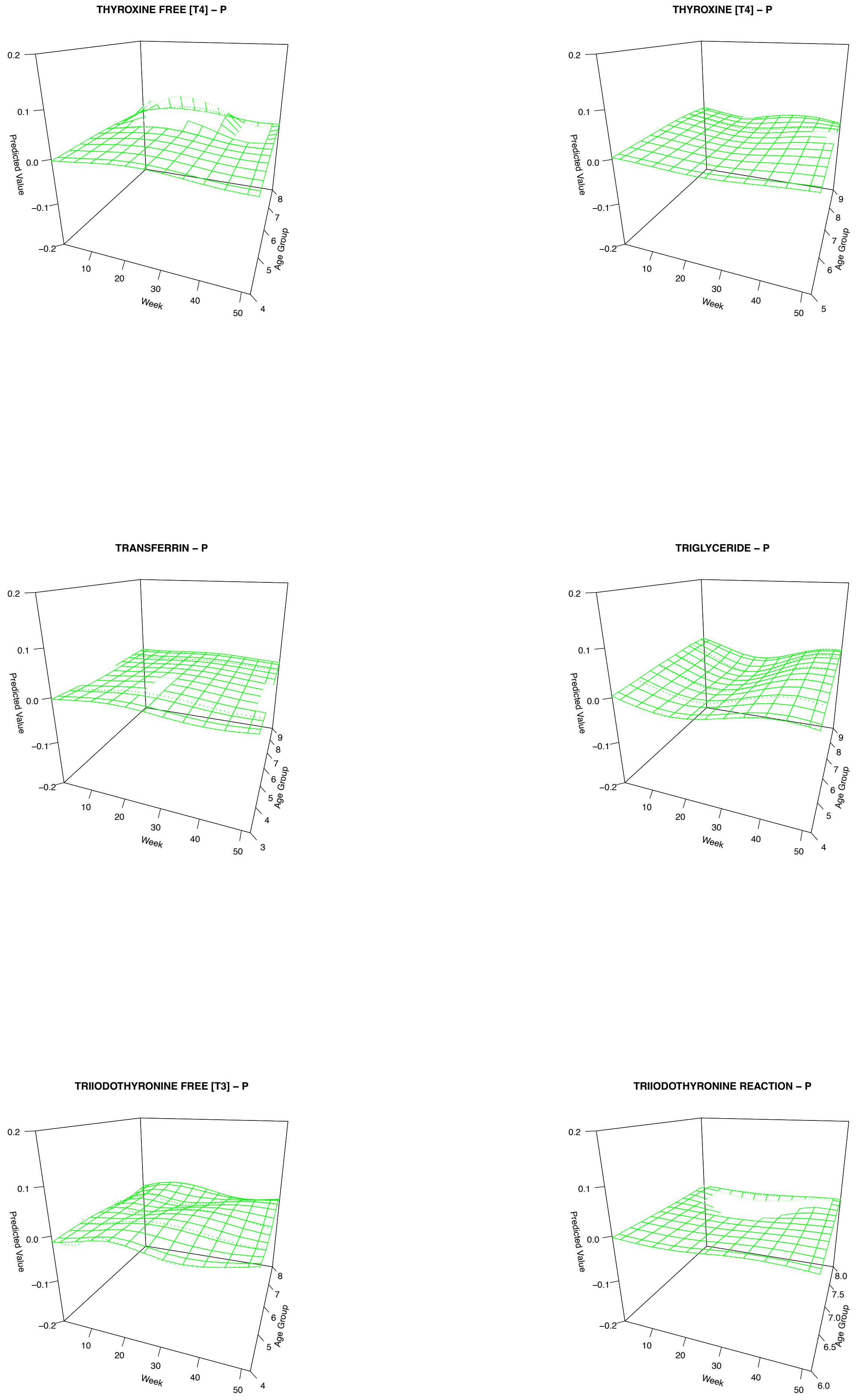

#### Supplemental Figure 2

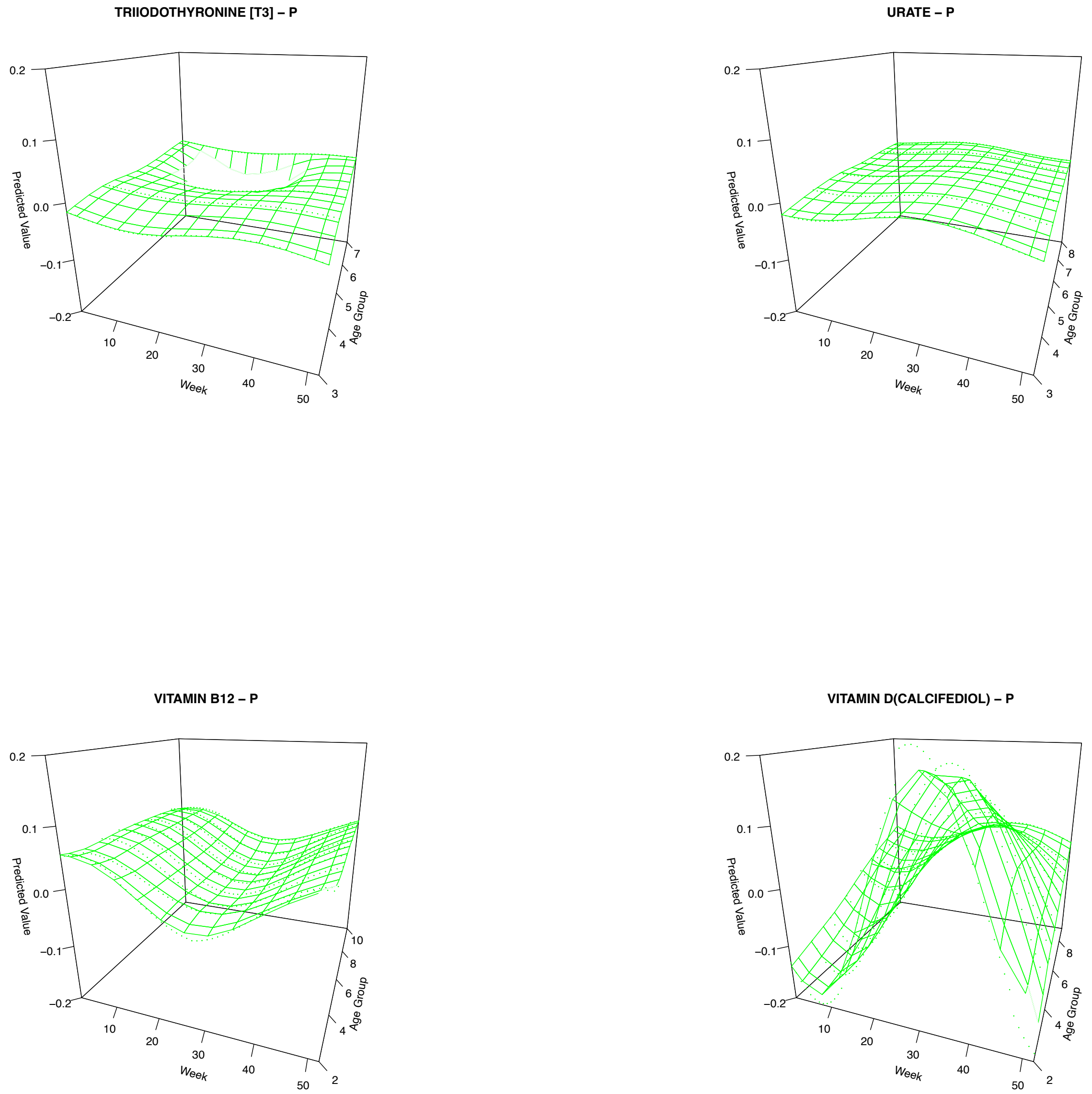


**Supplemental Figure 2. 3D mesh plots of predicted values by age group.** The figure displays 3D representations of predicted normalized values by age group, sex corrected. The x-axis represents week of the year where 1 corresponds to the first week of the year and 52 corresponds to the last week of the year. The y-axis shows the calculated predicted normalized value based on methods described in the paper. The z-axis shows the age group where the lowest age group correlates to the youngest cohort, while the larger age groups indicate older cohorts, also described in methods. Only unique tests with at least 3 significant age group fits were included in this figure.
